# Systematic review and meta-analyses on the prevalence of dementia in Latin America and Caribbean countries: Exploring sex, rurality, age, and education as possible determinants

**DOI:** 10.1101/2021.10.15.21265054

**Authors:** Fabiana Ribeiro, Ana C. Teixeira-Santos, Paulo Caramelli, Anja K. Leist

## Abstract

**Background:** Studies have shown that the prevalence of dementia in Latin America and the Caribbean (LAC) may be higher than in high-income countries. Thus, we sought to systematically analyse the prevalence of dementia and explore possible drivers that lead to this disparity in LAC countries.

**Method:** We searched Pubmed, Web of Knowledge, Scopus, Lilacs, and SciELO for studies on dementia in LAC countries in English, Spanish, and Portuguese languages. Random-effects model was applied.

**Results:** Twenty-nine studies from 12 LAC countries were included. Pooled prevalence of all-cause dementia was 11%. Further analyses with studies providing raw prevalence by sex, area, and educational level showed a higher prevalence for women (9%) than for men (5%). Also, dementia prevalence was higher for rural than urban residents (12% *vs* 8%, respectively). Participants without formal education presented more than double the prevalence of dementia (22%) compared to those with at least one year of formal education (10%). Studies with more recent data collection showed higher dementia prevalence.

**Conclusion:** Our findings suggest a high global dementia prevalence in LAC countries and an unequal burden of dementia for women, lower-educated, and rural residents. Secular raises in dementia prevalence call for increased public health efforts for preventative action.

**Highlights:**

- Rigorous and most comprehensive review of dementia prevalence in Latin America and the Caribbean (LAC) to date.
- Pooled dementia prevalence estimates are higher in LAC countries compared to high-income countries.
- Higher prevalence of dementia among women and lower-educated adults.
- Higher prevalence among older adults living in rural compared to urban areas.
- Qualitative analyses suggest a modest increase in dementia prevalence in LAC countries over time.

## Introduction

The population living in LAC countries was estimated to be around 653.962.000 people in 2020 (*UNdata*, 2020), accounting for 8.4% of the world population. Population ageing is one of the most crucial global transformations. In 1990, the world’s population aged 65 and over was 6%, and in LAC 5%, while by 2019, this number achieved 9% both worldwide and in LAC. Moreover, the proportion of people aged 65 years or older is expected to continue increasing so that one in six people in the world will be aged 65 or older in 2050. Consequently, more cases of dementia are expected to emerge (Custodio et al., 2017).

Dementia defines a range of chronic disorders characterised by cognitive and functional impairment that comprise a variety of specific medical conditions, e.g., vascular dementia, dementia with Lewy bodies, frontotemporal dementia, and Alzheimer’s disease; the latter corresponds to 60-70% of cases (World Health Organization, 2020). The Global Burden of Disease Study, using 2016 global data, found that the number of individuals living with Alzheimer’s disease and other dementias increased 117% from 1990 to 2016, with a higher prevalence among women compared with men. Moreover, dementia was the fifth leading cause of death worldwide and consistently ranked among the top three causes of disability in most countries (GBD 2016 Dementia Collaborators, 2019).

Dementia is one of the most significant causes of dependence among older adults, causing considerable medical and care costs (Custodio et al., 2017; Parra et al., 2018; Prince et al., 2015), which counts to 0.2% of the total gross domestic product in low- and middle-income countries and 1.4% in rich countries (World Health Organization, 2020). Thus, increases in the prevalence of dementia will boost the difficulties in public health systems of LAC countries, increasing the already high economic and social burden related to dementia in this region of the world (Ibáñez et al., 2021), implying significant efforts to adapt social and public policies accordingly (Prince et al., 2013). Further, dementia prevalence in older age signals the capacity of older adults for financial assistance and support towards younger family members (e.g., through caregiving for grandchildren). In LAC countries, older adults substantially contribute to family income and care when they are healthy but depend on the support of younger family members when they have a compromised health condition (Gomes, 2007; Pelaez & Martinez, 2002). For this reason, it is essential to evaluate and systematise studies of the prevalence of dementia in LAC countries, taking into account possible differences among countries, which would help them plan for tailored preventive measures and long-term care policies (Custodio et al., 2017; Mukadam et al., 2019).

A previous review performed by Nitrini et al. (2009) assessed the prevalence of dementia in older adults (65 to 90+ years) in Latin America based on eight studies and found no differences in the prevalence compared to developed countries (7.1%). Nevertheless, they pointed out that those participants aged 65 to 69 years had a higher dementia prevalence, possibly related to the higher number of illiterates among those individuals and to a poor control of cardiovascular risk factors.

A more recent systematic review explored the prevalence of dementia globally between 1980 and 2009, showing that age-standardised prevalence for those aged 60+ years old living in Latin America (11 studies) was higher (8.5%) compared to other world regions, i.e., Europe (6.9%), North America (6.5%), or Asia (6.9%). In their review, dementia prevalence increased with age and was higher in women than men, with increasing sex gaps with advancing age (Prince et al., 2013). These sex effects were explained by increased survival in women with dementia, as well as generally increased incidence of dementia among women and men (Brodaty et al., 2012; Carter et al., 2012). Although this study included diverse authors fluent in different languages, their bibliographical search strategy only included terms in English.

A recent meta-analysis carried out by Cao et al. (2020), including articles in English and Chinese, investigated dementia prevalence in Asia, Africa, Latin America, and Europe/North America. From the 47 selected articles with sex-specific prevalence, only two studies were from Latin America, making valid inferences about dementia prevalence in this world region difficult.

Finally, two systematic reviews specifically exploring the prevalence of dementia in LAC countries revealed discrepant prevalence rates. Zurique Sánchez et al. (2019) included 25 articles published in Spanish and English in their review. They found a general prevalence of 11% with a higher dementia prevalence in women compared to men, and in rural samples compared to urban ones. However, this review did not have strict criteria for the diagnosis of dementia. Instead, the authors included studies in which the dementia diagnosis was based only on screening tests, such as the Mini-Mental State Examination (MMSE). On the other hand, Xiang et al. (2021) used more strict criteria for dementia diagnosis and found a lower prevalence (8%). However, only seven studies, published in English between 2013 and 2018, were included in this review.

As previous systematic reviews on the prevalence of dementia in the older adult population in LAC countries provide limited evidence, this study aimed to deliver a more integrative analysis of the current literature of the prevalence of dementia in LAC countries, strategically searching for articles published in English or in the main official languages of this region, as well as precise estimates of all-cause dementia prevalence, by sex and area (rural and urban), in addition to qualitatively analysing the influence of age, education, and year of data collection on dementia prevalence.

## Method

We followed the PRISMA guidelines by Moher et al. (2010) and the recommendations of the Cochrane Collaborations for this systematic review and meta-analysis.

### Literature search

We carried out a systematic literature search for studies assessing the prevalence of dementia in adults aged 50 or more in LAC countries in five databases: PubMed, Web of Science, Scopus, Lilacs, and SciELO. On August 16, 2021, the search was performed in three languages: English, Portuguese, and Spanish, without date restriction. The search terms used in each database are provided in the Supplementary Materials. We also examined the reference list of previous systematic reviews assessing the prevalence of dementia in Latin America (Cao et al., 2020; Nitrini et al., 2009; Prince et al., 2013; Xiang et al., 2021; Zurique Sánchez et al., 2019).

### Criteria for inclusion and exclusion of studies

The inclusion criteria for articles to be selected in this systematic review were as following: i) cohort or cross-sectional study designs reporting population- or community-based data, from population surveys or patients identified in representative samples of LAC countries; ii) population-based studies, including the prevalence of dementia in participants aged ≥ 50; and iii) Studies including frequency of any type of dementia with clearly defined diagnostic criteria, such as with Diagnostic and Statistical Manual of Mental Disorders (DSM), International Statistical Classification of Diseases and Related Health Problems 10th revision (ICD-10), and/or clinical interview performed by trained professionals.

The exclusion criteria of the articles were applied as follows: i) duplicated studies; ii) articles including only samples with “Mild cognitive impairment”, “cognitive impairment”, or “cognitive impairment no dementia”; iii) articles focused on sample populations outside Latin America and Caribbean, and iv) studies including secondary or tertiary care services in hospital or clinical-based samples were also excluded to avoid selection bias.

### Data extraction and assessment of methodological quality

A standard data form was developed to draw the main estimates from the selected studies. FR extracted data in an Excel spreadsheet for each article, and ACT reviewed the spreadsheet.

The majority of studies only provided diagnostic assessments of all-cause dementia, few studies reported assessment of subtypes of dementia (*n* = 9), and just one specifically assessed Alzheimer’s disease. We, thus, only present prevalence estimates of all-type dementia. Figures and estimates collected from each study include the total, number and percentage by sex of respondents with all-type dementia or/and subtypes when provided (e.g., Alzheimer’s disease, vascular dementia, dementia with Lewy bodies); diagnostic criteria and screening instruments for dementia; study design; prevalence rates; essential sociodemographic characteristics when available; such as total sample size, age, sex, and education; and information regarding the rurality of the sample and the socioeconomic status of participants indicated by income.

The methodological quality of the selected studies was assessed by FR and reviewed by ACT through the JBI Critical Appraisal Checklist for Studies Reporting Prevalence Data (Munn et al., 2015), a 9-item checklist designed to evaluate the methodological quality of the studies and to establish possible bias in the design, conduct, and analysis. Any inconsistencies among reviewers about the quality of the articles were resolved by consensus.

### Data analysis

In order to estimate the pooled prevalence of all-cause dementia, the rate of participants diagnosed with dementia by sample was included in our first analysis. In addition, we also run separate study-pooled prevalence of all-cause dementia by sex, area (urban *vs* rural), and educational level (no formal education *vs* at least one year of formal education). In this meta-analysis, participants without formal education and illiterate participants were categorized into one group ‘no formal education’.

Prevalence with a 95% confidence interval was assessed via the inverse variance method. Statistical heterogeneity was evaluated using I^2,^ in which values ≥ 75% were considered an indicator of substantial heterogeneity. Although only studies that used strict diagnostic criteria for dementia assessment were included, high heterogeneity was expected, in line with previous meta-analyses (Bacigalupo et al., 2018; Cao et al., 2020; Zurique Sánchez et al., 2019). Accordingly, the random effects method was used to estimate the pooled prevalence.

Sensitivity analyses were performed by removing studies in which dementia was based on one-phase diagnostic assessment to explore possible bias from one-versus two-phase assessment. The leave-one-out method was also performed to examine the robustness of the findings for the pooled prevalence of all-cause dementia. Finally, we empirically checked publication bias by applying Egger’s regression test and visually creating funnel plots (provided in Supplementary Materials). All statistical analyses were carried out in STATA 17, using the METAPROP command (StataCorp, 2021).

## Results

### Study selection process

The database search yielded 3395 citations, and two additional articles were found in the reference list of Nitrini et al. (2009), resulting in a total of 3397 records. After importing them into Zotero, duplicated studies were filtered. The remaining 2693 citations were independently screened by title and abstract by two independent reviewers (FR and ACT). Following this, 2693 studies were excluded, and 228 articles were selected for full-text screening. There was a good Kappa inter-rater reliability agreement between reviewers over all the records screened *(n* = 2693), including both included and excluded studies (*k* = 0.75) (Fleiss, 1981). The most frequent reasons for exclusion during the full-text screening phase were the study design (not a population- or community-based sampling) (*n* = 71) or lack of strict criteria for dementia diagnosis (*n* = 30). Following the full-text, 32 articles were selected and included in the qualitative analysis, while 29 comprised the quantitative analysis. The flow chart of the study selection process and the reason for excluding publication is provided in Figure 1.

**Figure 1.**
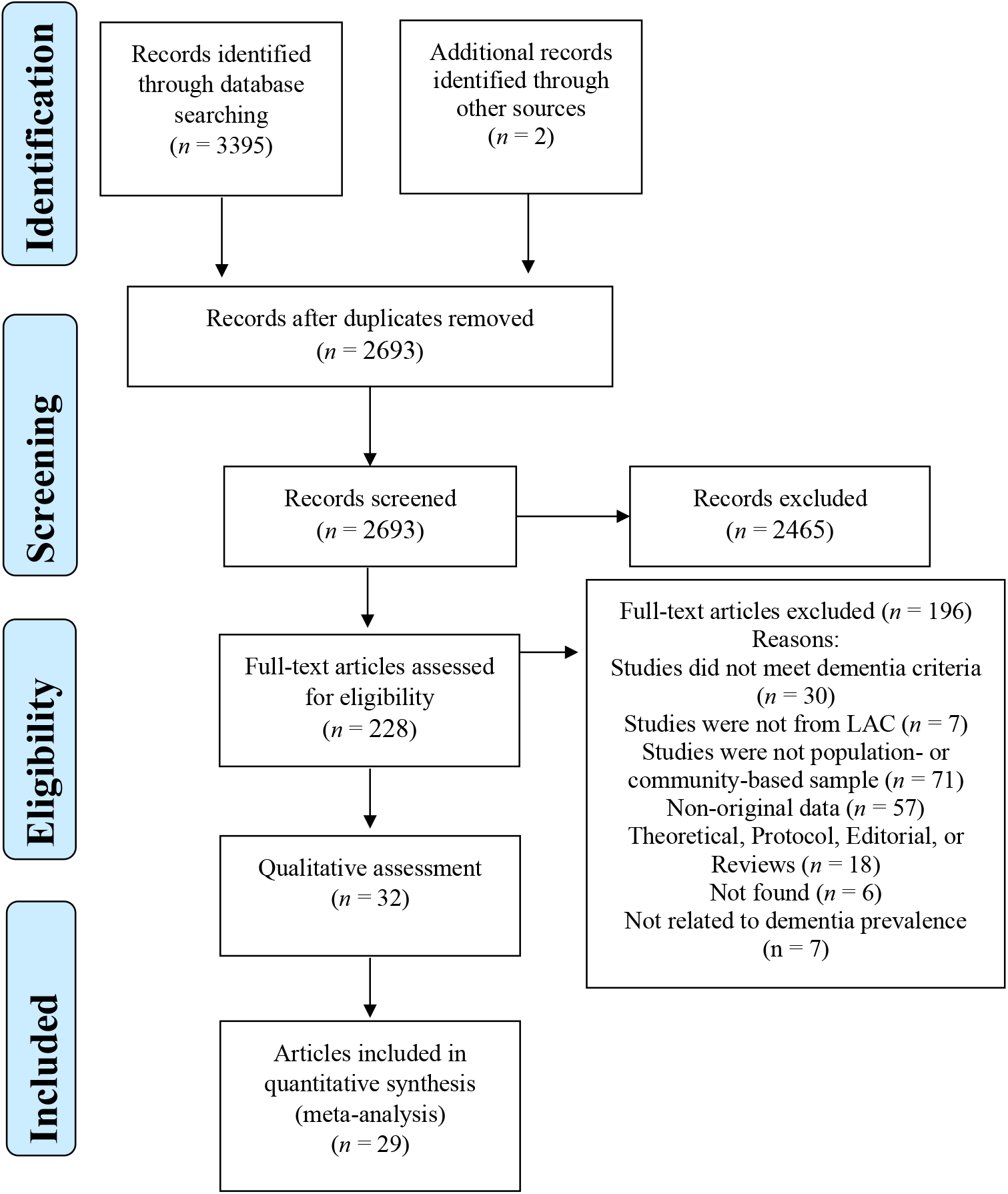
Flow diagram of the study selection process.

### Characteristics of the selected studies

Table 1 sums up the characteristics of the 31 articles and a conference abstract included in this systematic review. Twenty-four articles and the abstract were published in English, and the remaining seven were published in Spanish. In total, they comprise 37 different samples published between 1997 and 2021. Sample sizes ranged from 101 to 18,351 respondents, which corresponds to 91,796 participants in total. Fourteen of the 20 LAC countries were covered: Argentina, Brazil, Chile, Colombia, Cuba, Dominican Republic, Honduras, Jamaica, Mexico, Panama, Peru, Trinidad and Tobago, Uruguay, and Venezuela in the qualitative analyses.

**Table 1.**
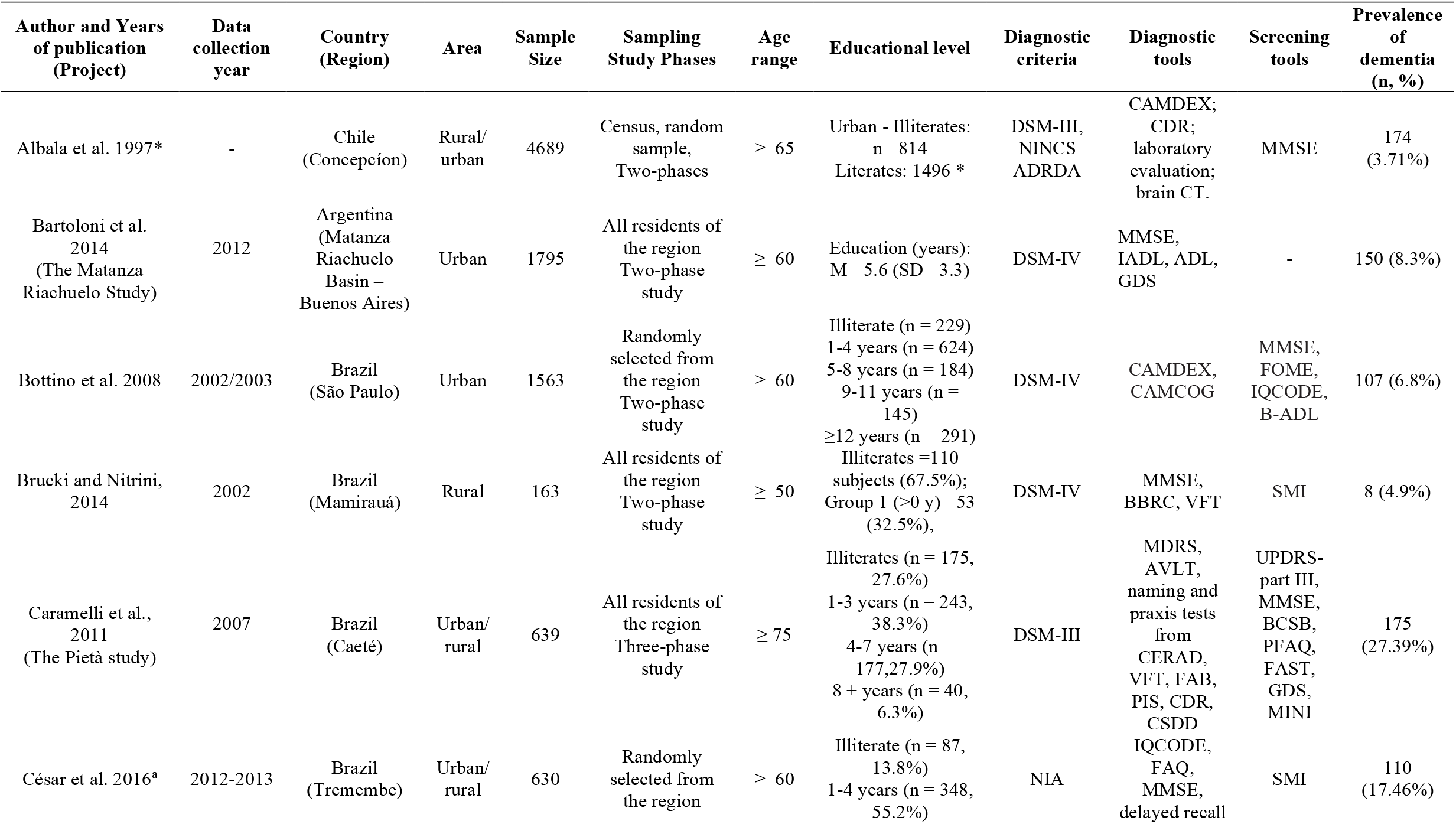
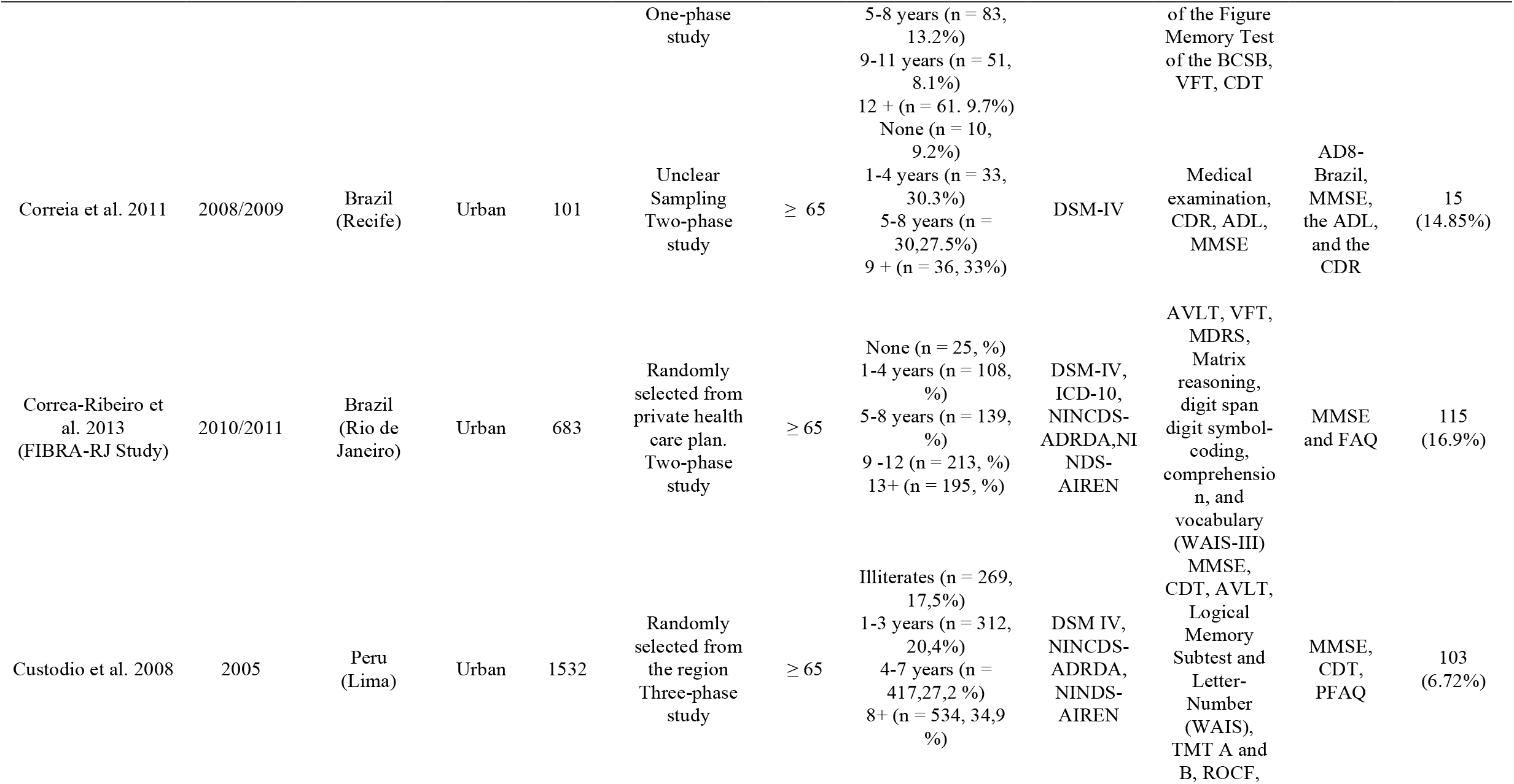
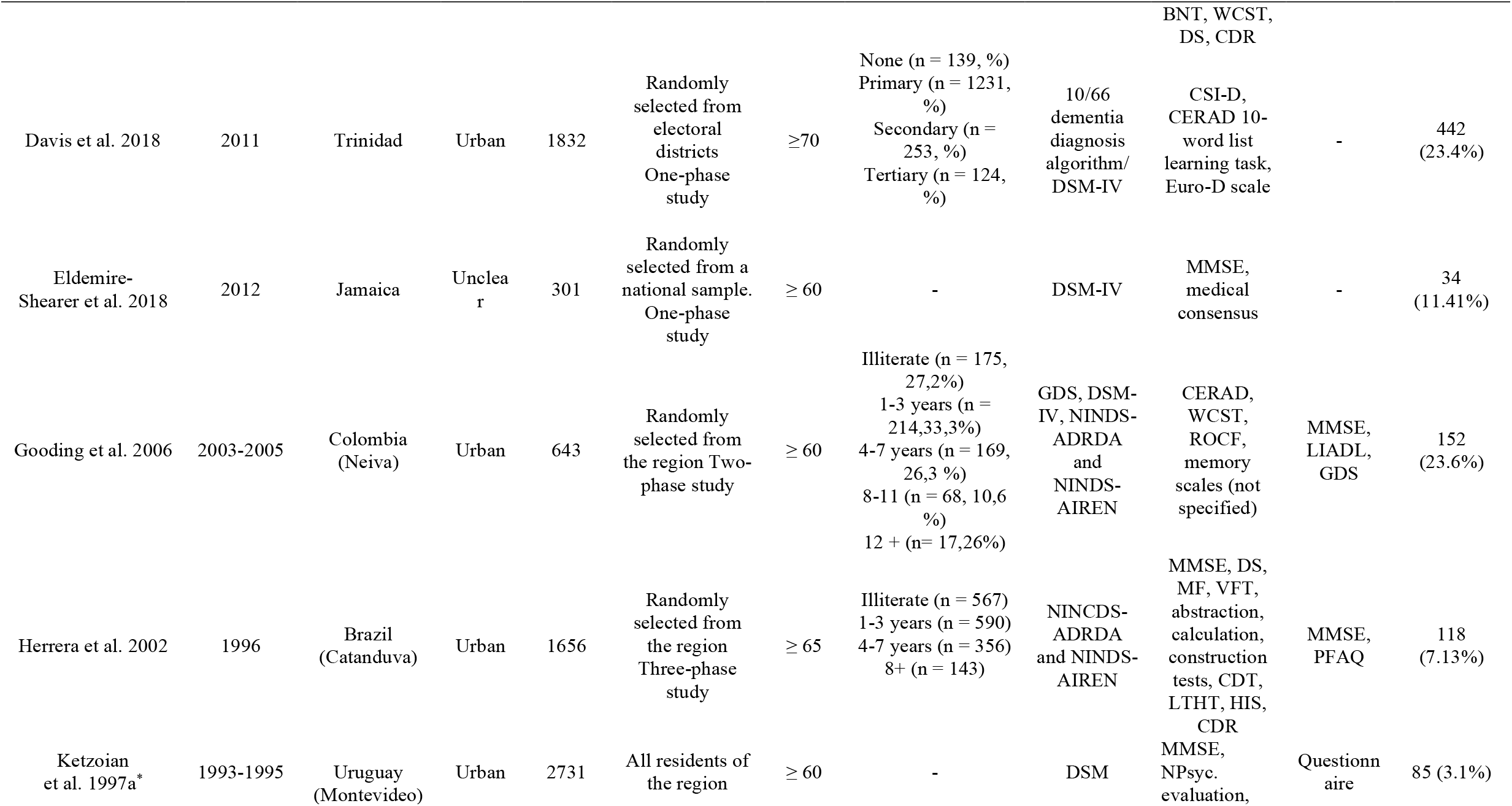
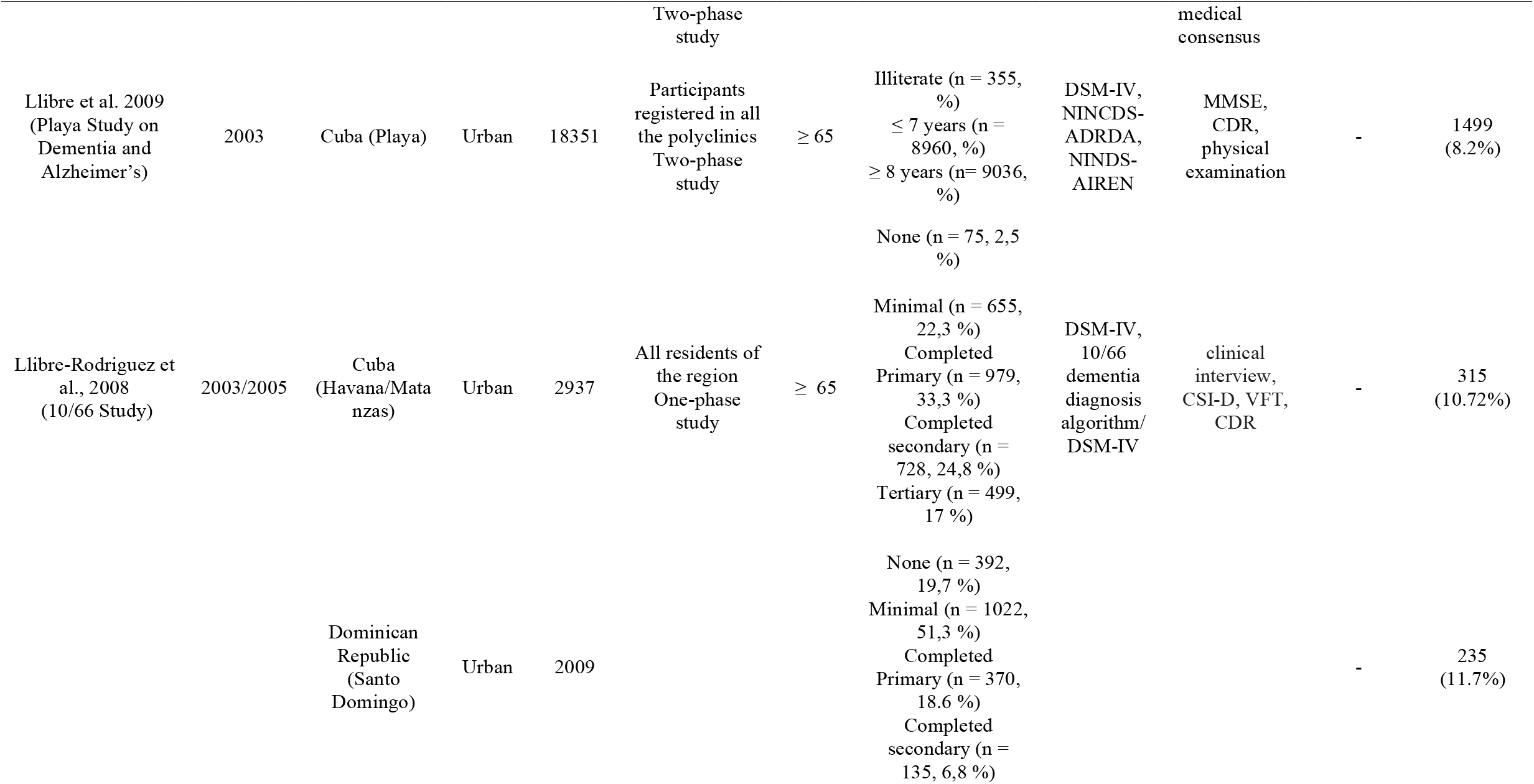
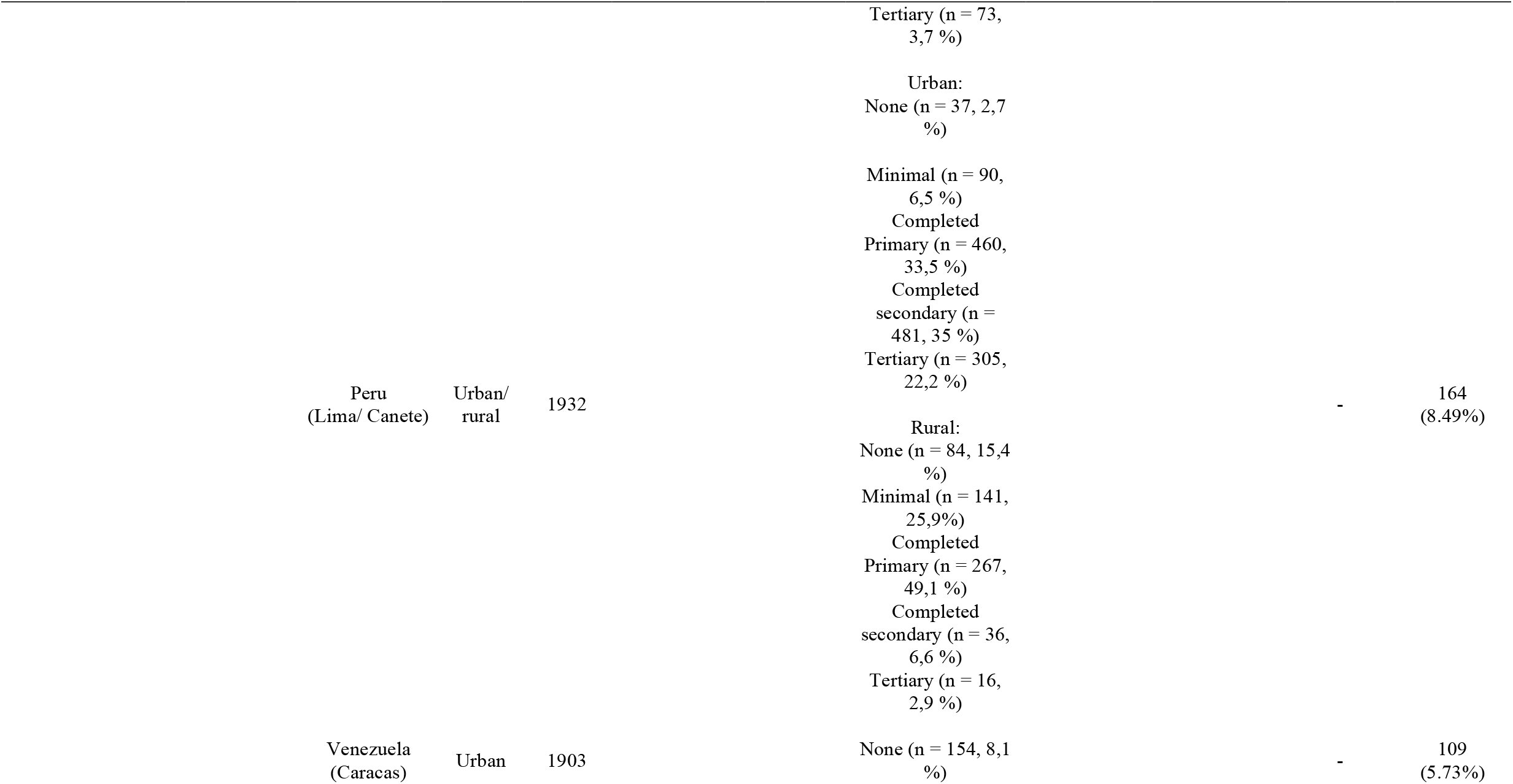
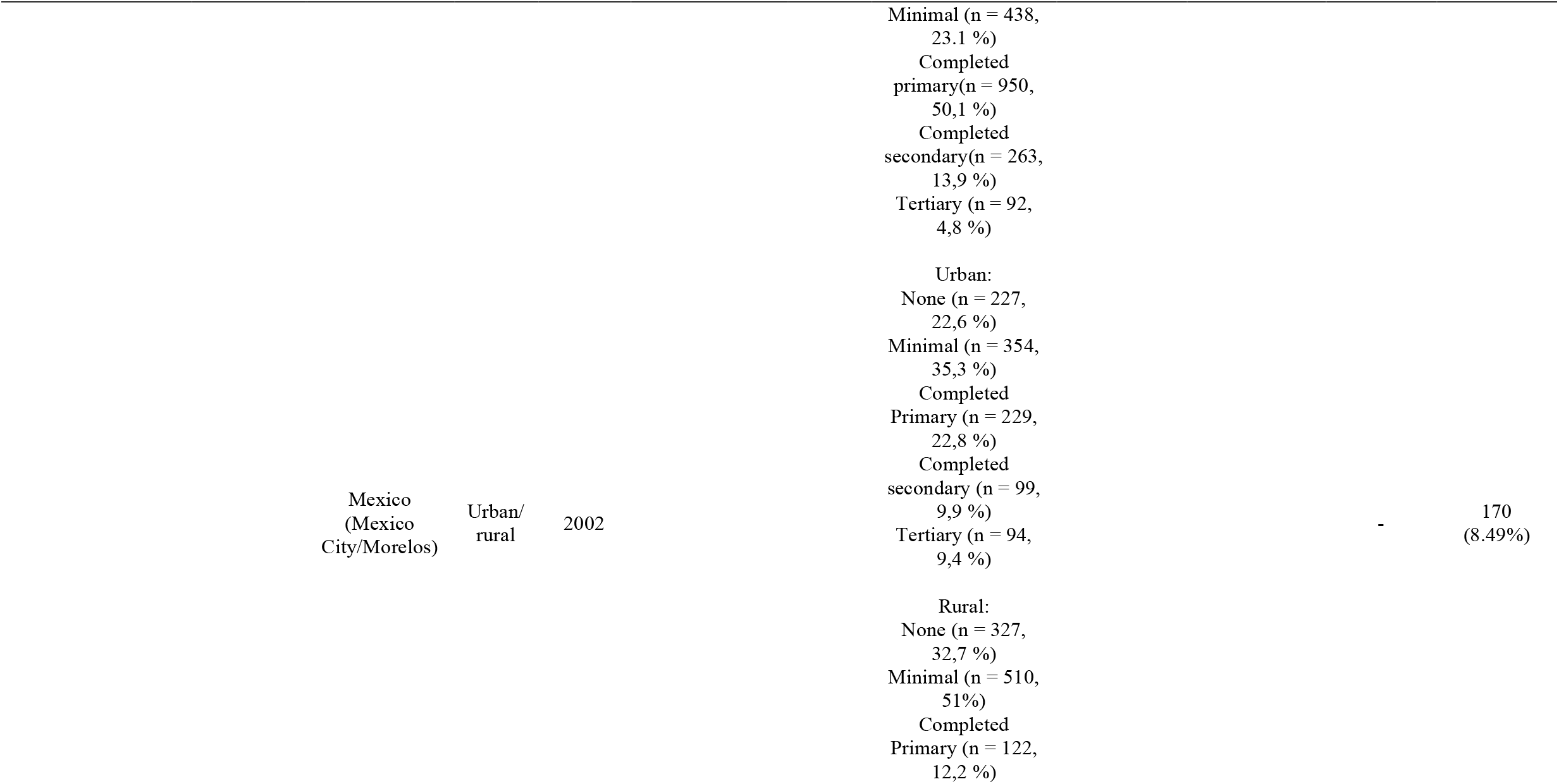
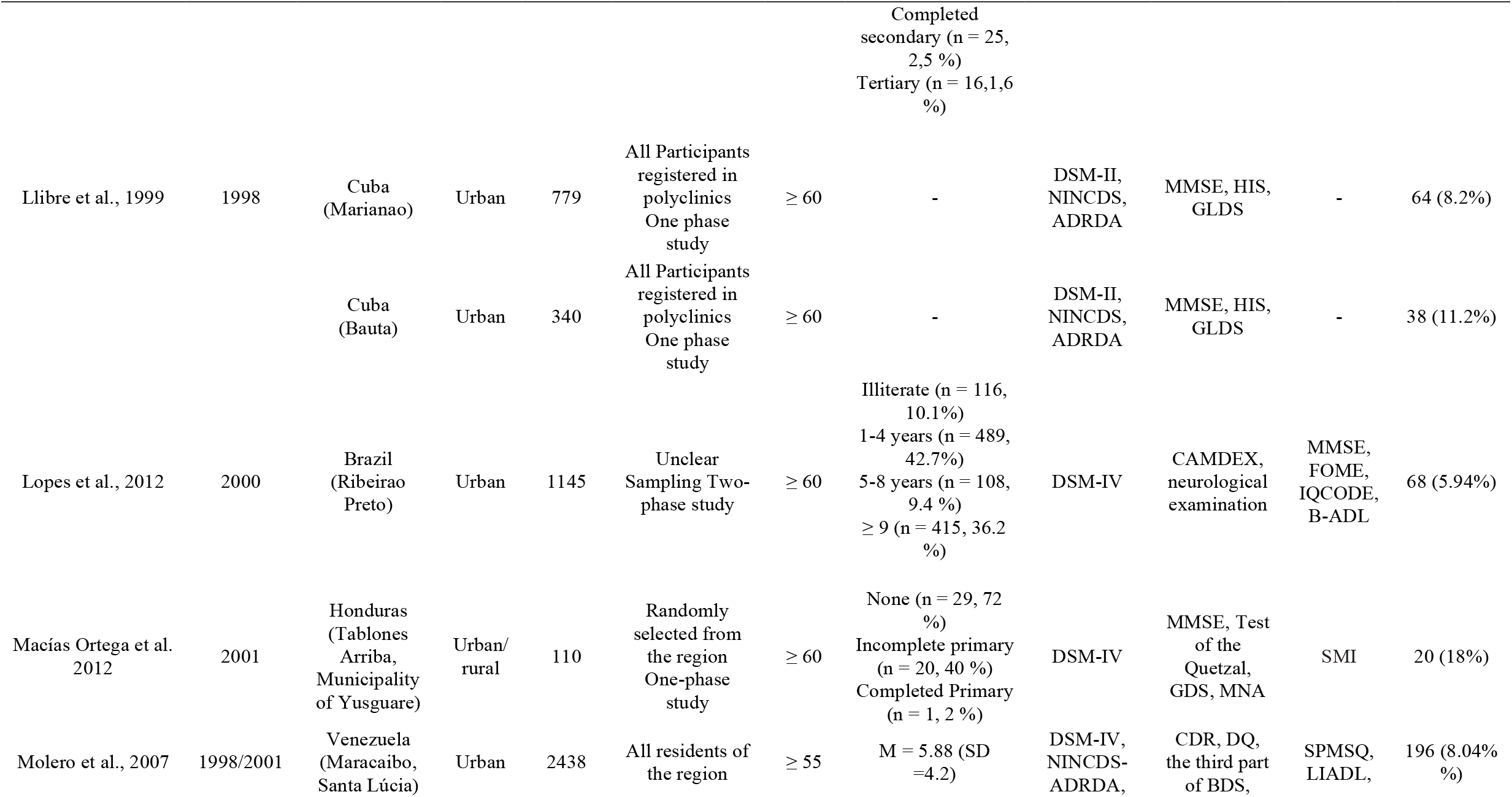
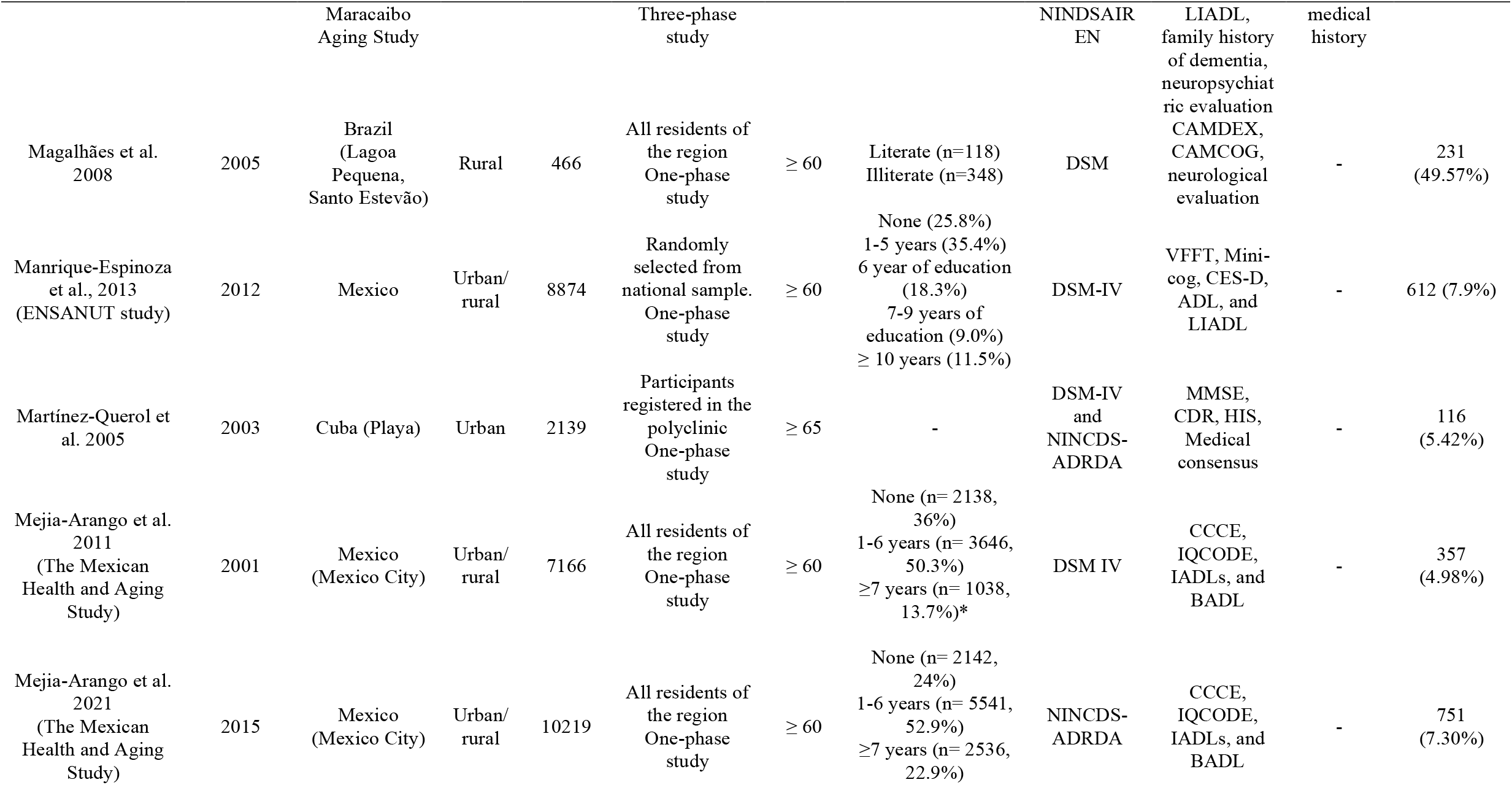
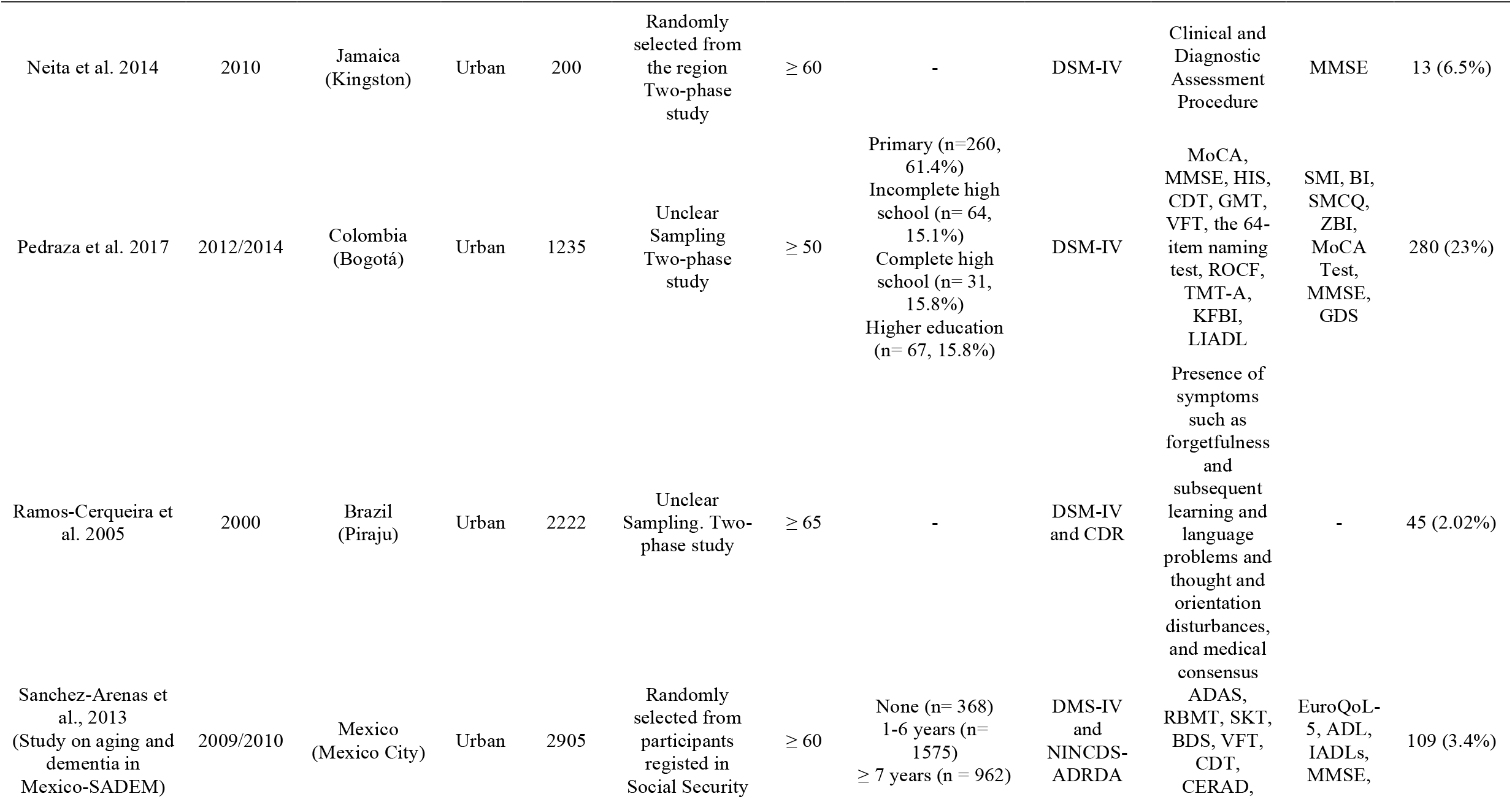
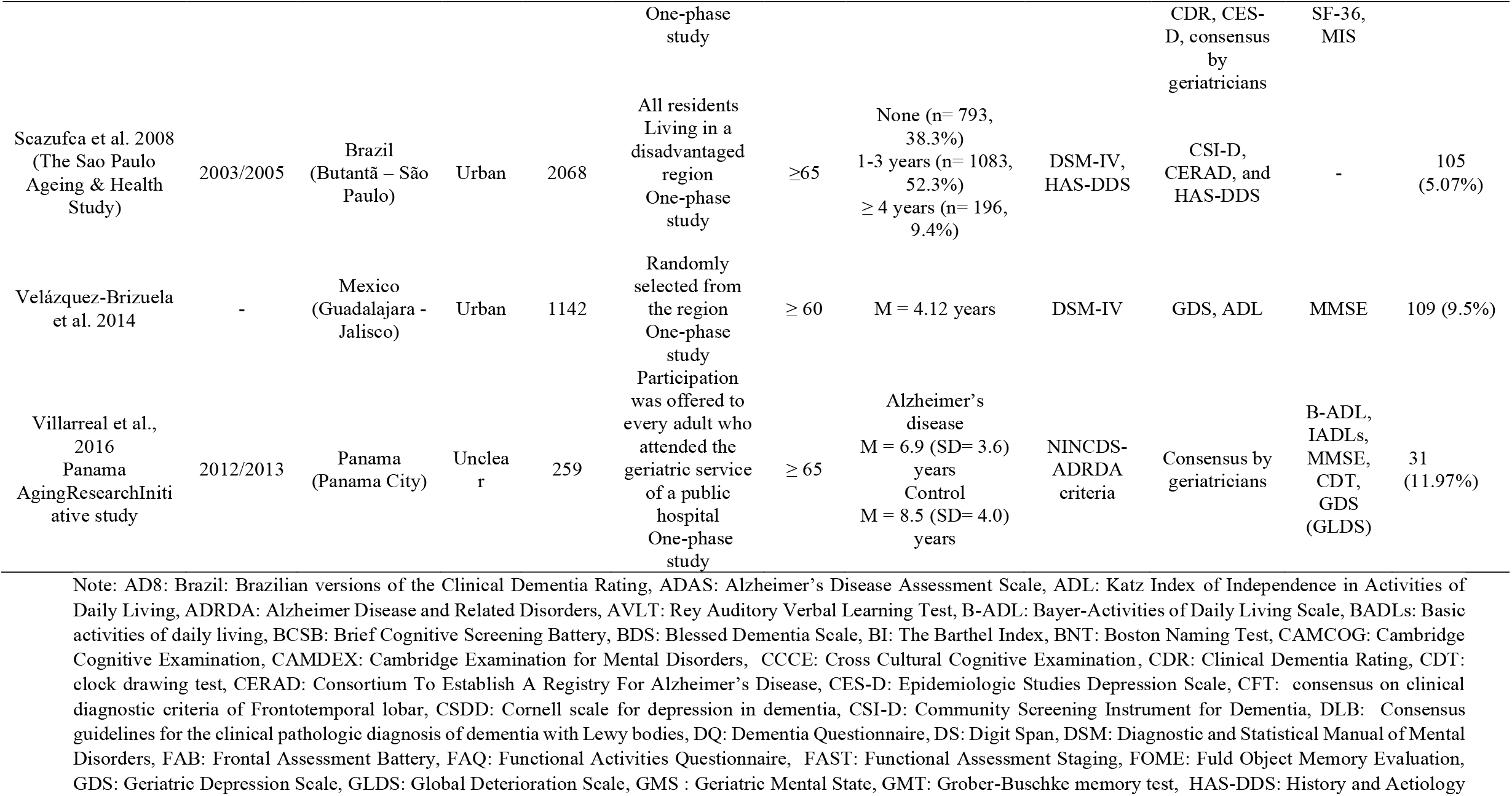
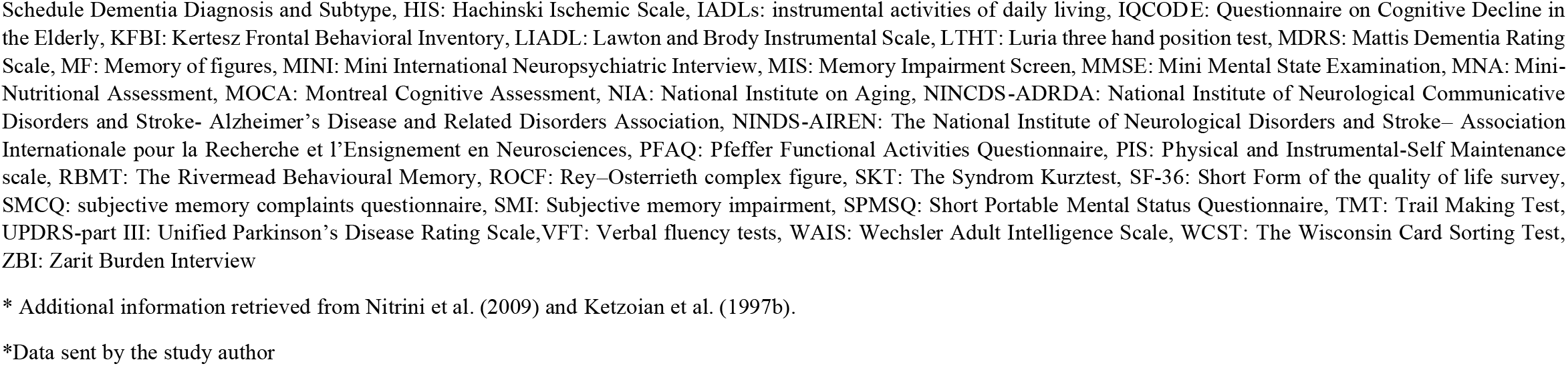
Characteristics of selected studies.

Regarding rurality, most of the studies included participants in urban areas (*n* = 24), while two studies from Brazil comprised individuals from rural populations, and nine studies had individuals recruited in both, rural and urban areas.

A total of nine articles specified the types of dementia, such as Alzheimer’s disease and vascular dementia (Bottino et al., 2008; Correa Ribeiro et al., 2013; Custodio et al., 2008; Herrera et al., 2002; Llibre Rodríguez et al., 1999; Maestre et al., 2018; Martínez Querol et al., 2005; Scazufca et al., 2008; Villarreal et al., 2016).

Studies differed on whether they assessed dementia in one phase, with a one-time full assessment or two or more phases, with an initial screening phase and subsequent in-depth assessment of participants with probable dementia (the pooled prevalence by the number of diagnostic phases is provided in supplementary material Figure S1). Thirteen out of the 32 included studies conducted a two phases-diagnosis (Albala et al., 1997; Bartoloni et al., 2014; Bottino et al., 2008; Brucki & Nitrini, 2014; Correa Ribeiro et al., 2013; Correia et al., 2011; Gooding et al., 2006; Ketzoian et al., 1997; Llibre et al., 2009; Lopes et al., 2012; Neita et al., 2014; Pedraza et al., 2017; Ramos-Cerqueira et al., 2005) and four studies conducted more than two phases-diagnosis to identify patients with dementia (Caramelli et al., 2011; Custodio et al., 2008; Herrera et al., 2002; Molero et al., 2007). In general, MMSE was used as the main instrument to screen symptoms of dementia; however, the following phases varied widely among studies in terms of instruments used.

As provided in Table 1, the Diagnostic and Statistical Manual of Mental Disorders (DSM-III, DSM-IV, or DSM-V) was the most used criteria for diagnosing dementia. However, some studies used the 10/66 algorithm in addition to the DSM criteria (Davis et al., 2018; Llibre-Rodriguez et al., 2008). Besides the DSM, one study used the History and Aetiology Schedule Dementia Diagnosis and Subtype (Scazufca et al., 2008), another one applied the National Institute on Aging criteria (César et al., 2016), and one of them used the cut-off points of the Cambridge Examination for Mental Disorders (Magalhães et al., 2008).

Some studies aimed specifically to diagnose Alzheimer’s disease, and in this case, the search revealed nine studies using the criteria established by the National Institute of Neurological and Communication Disorders and Stroke and the Alzheimer’s Disease and Related Disorders Association (NINCDS-ADRDA) (Albala et al., 1997, Correa-Ribeiro et al., 2013, Custodio et al. 2008, Gooding et al. 2006, Herrera et al. 2002, Llibre et al. 2009, Llibre et al., 1999, Molero et al., 2007, Martínez et al. 2005, Sanchez-Arenas et al., 2013, Villareal et al., 2016). In the case of vascular dementia, the diagnosis was established based on criteria from the National Institute of Neurological Disorders and Stroke Association Internationale pour la Recherche et l’Enseignement en Neurosciences (NINDS-AIREN) (Correa-Ribeiro et al., 2013; Custodio et al. 2008, Gooding et al. 2006, Herrera et al. 2002, Llibre et al. 2009, Molero et al., 2007). As Villarreal et al. (2016) assessed only Alzheimer’s Disease, we excluded it from the quantitative analyses of all-cause dementia.

### Methodological quality of the included studies

The methodological rigour of each study was assessed by applying the nine criteria provided by the JBI critical appraisal checklist for studies reporting prevalence data. Only studies meeting a minimum of five criteria were included in the quantitative analysis (see supplementary material, Table S1). Furthermore, one of the criteria was related to the dementia diagnosis assessment, which should consist of a valid method for identifying dementia. This was a mandatory criterion in itself for a study to be included in this systematic review. Specifically, eight articles had total scores on the JBI critical appraisal checklist (Bottino et al., 2008; César et al., 2016; Custodio et al., 2008; Davis et al., 2018; Llibre et al., 2009; Llibre Rodriguez et al., 2008; Molero et al., 2007; Scazufca et al., 2008),

All but four studies (Bartoloni et al., 2014; Correa Ribeiro et al., 2013; Pedraza et al., 2017; Villarreal et al., 2016) did not include representative sample frames to address the target population. Bartoloni et al. (2014) excluded illiterate participants from the final sample, limiting the sample’s representativeness, since the illiteracy rate is high in older adults in LAC countries (UNESCO, 2017). Correa Ribeiro et al. (2013) only included participants from a private health care plan, which represented less than 30% of the older adults in Rio de Janeiro at the time of data collection (Agência Nacional de Saúde Suplementar, 2012). The description of the study methods by Pedraza et al. (2017) did not conclusively state whether the sample was representative, and Villarreal et al. (2016) comprised older adults of just one public hospital in Panama, for this reason, their results may underestimate the extent of dementia among older Panamanians (Villarreal et al., 2019). Although five articles were classified as unclear regarding the appropriateness of recruited participants (Brucki & Nitrini, 2014; Correia et al., 2011; Lopes et al., 2012; Pedraza et al., 2017; Ramos-Cerqueira et al., 2005), all articles presented adequate sample size. Moreover, in 11 studies, it was not clear whether dementia was assessed in a standardised way, i.e., whether those involved in data collection were trained in the use of the instruments (Albala et al. 1997; Eldemire-Shearer et al. 2018; Macías Ortega et al., 2012; Magalhães et al., 2008; Martínez Querol et al., 2005; Mejia-Arango & Gutierrez, 2011; Mejia-Arango et al., 2021; Neita et al., 2014; Pedraza et al., 2017; Sanchez-Arenas et al., 2013; Villarreal et al., 2016).

Regarding the appropriateness of the statistical analysis, 14 articles did not report confidence intervals (Albala et al. 1997; Bartoloni et al., 2014; Brucki & Nitrini, 2014; Caramelli et al., 2011; Eldemire-Shearer et al. 2018; Gooding et al., 2006; Herrera et al., 2002; Ketzoian et al. 1997a; Macías Ortega et al., 2012; Manrique-Espinoza et al., 2013; Martínez Querol et al., 2005; Neita et al., 2014; Ramos-Cerqueira et al., 2005; Villarreal et al., 2016).

Finally, we excluded from quantitative analyses two articles that did not present valid methods for identifying dementia. In one study, it was unclear if there was a clinical assessment (Macías Ortega et al., 2012). In the other study, only MMSE cut-off scores were used to diagnose dementia, although they reported using the DSM diagnostic criteria (Velázquez-Brizuela et al., 2014).

### Quantitative analyses

As displayed in Figure 2, the pooled prevalence of all-cause dementia throughout the 30 included studies was 11% (95% CI: 8% - 14%). We detected statistically significant heterogeneity between studies (I^2^= 94.44%, *p* < 0,001) and significant publication bias by Egger’s test (t = 2.12, *p* = 0.04) (the funnel plot is available as Figure S1), which pointed to the need for further analyses to explore the source of heterogeneity.

**Figure 2.**
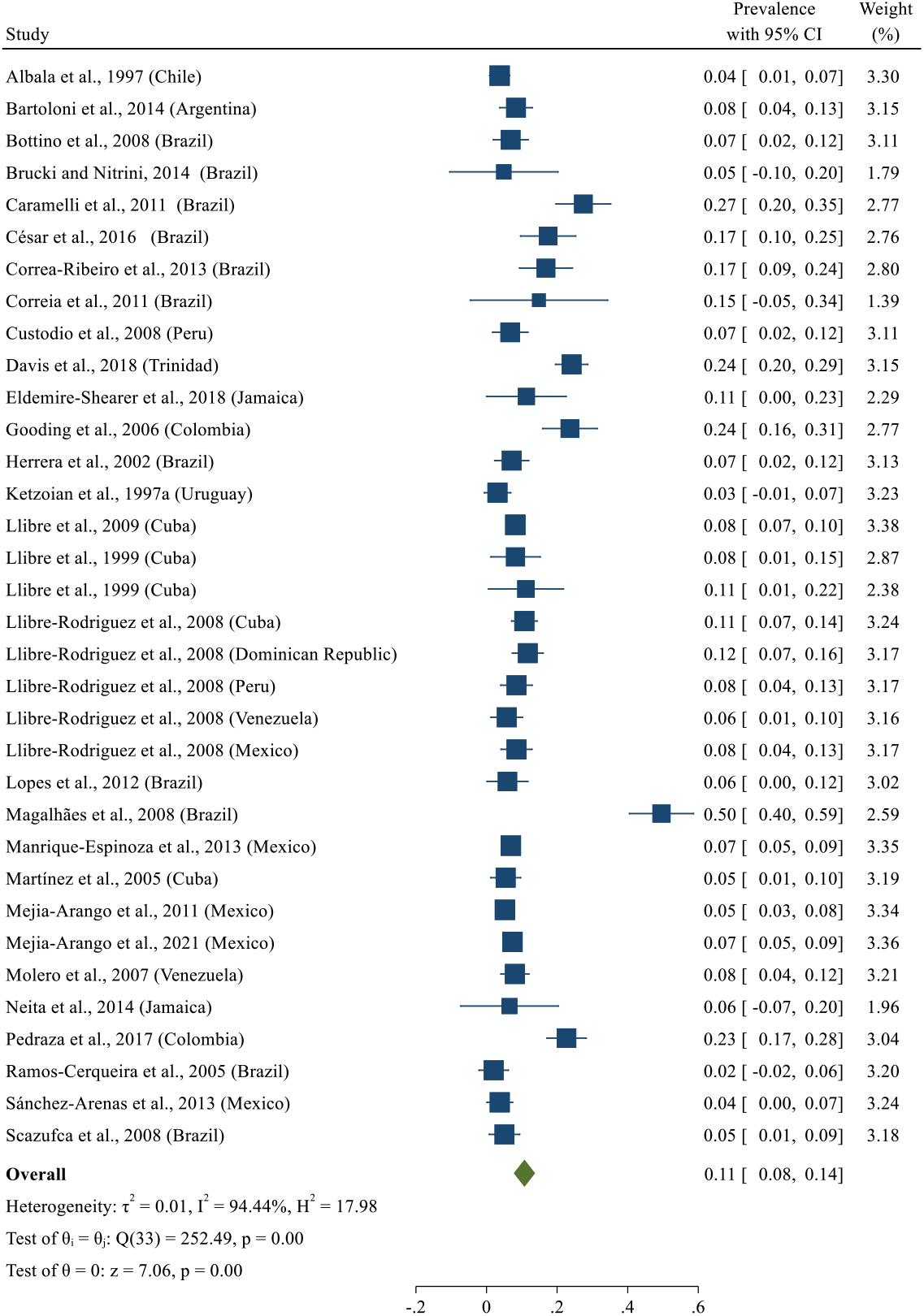
All-cause dementia prevalence.

In a second step, the prevalence of all-cause dementia was pooled after excluding the studies with one-phase diagnosis. A prevalence of 10% (95% CI: 6% - 14%) was observed (Figure S2), with no publication bias (*t* = 1.16, *p* = 0.24) (for the funnel plot, see Figure S3).

Moreover, we performed an analysis including only studies with representative samples; for this, three studies were excluded (Bartoloni et al., 2014; Correa Ribeiro et al., 2013; Pedraza et al., 2017), and the prevalence was 10% (95% CI: 7% - 13%) (Figure 3).

**Figure 3.**
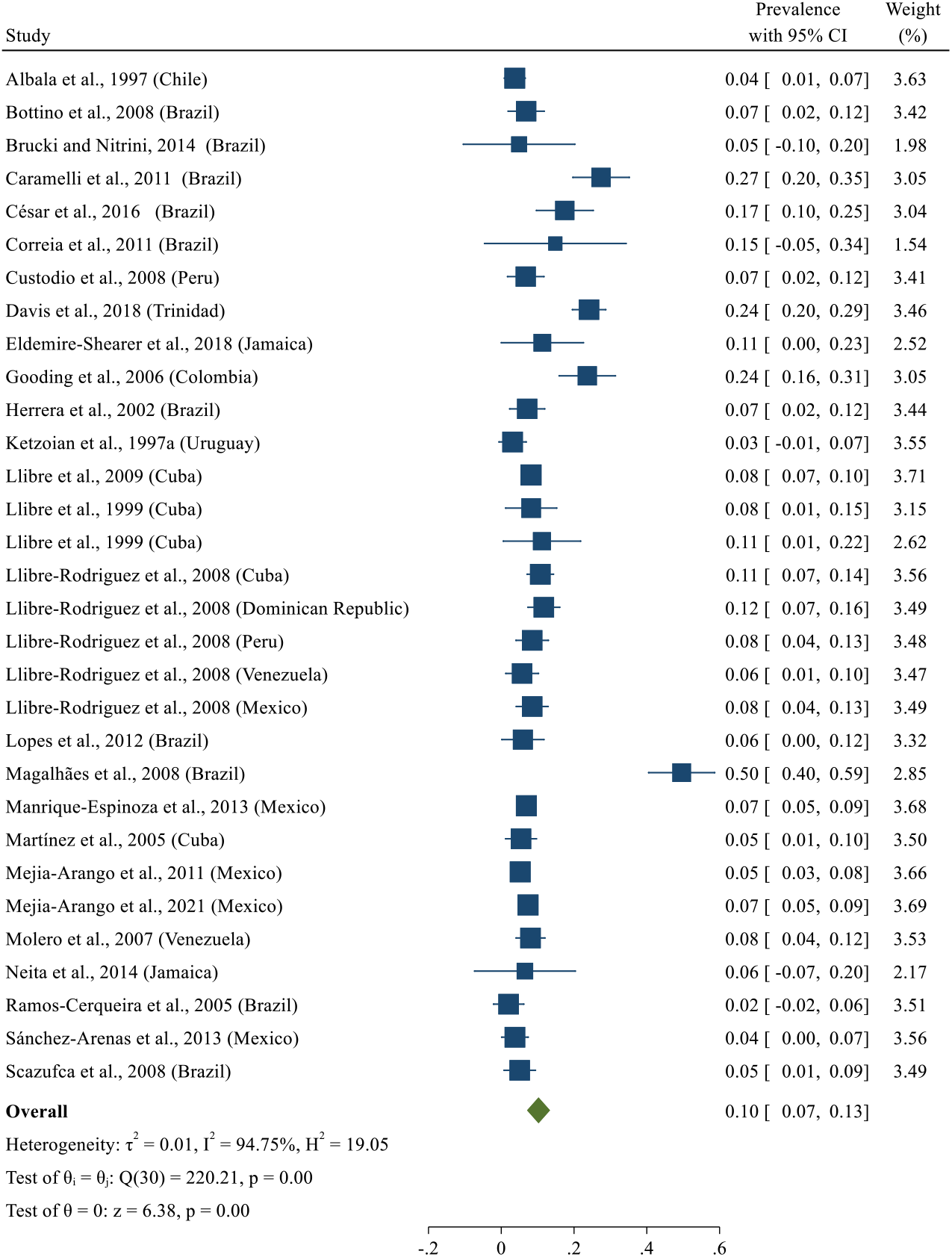
All-cause dementia prevalence for studies with representative samples.

We also carried out the leave-one-out method (see Figure S4), to observe if any of the studies was influencing the prevalence. Here, only Magalhães et al. (2008) study increased the pooled prevalence of all-cause dementia; the difference between this study and the remaining ones is that the former only included participants from rural areas. After excluding this article, the pooled prevalence decreased to 9% (see Figure 4).

**Figure 4.**
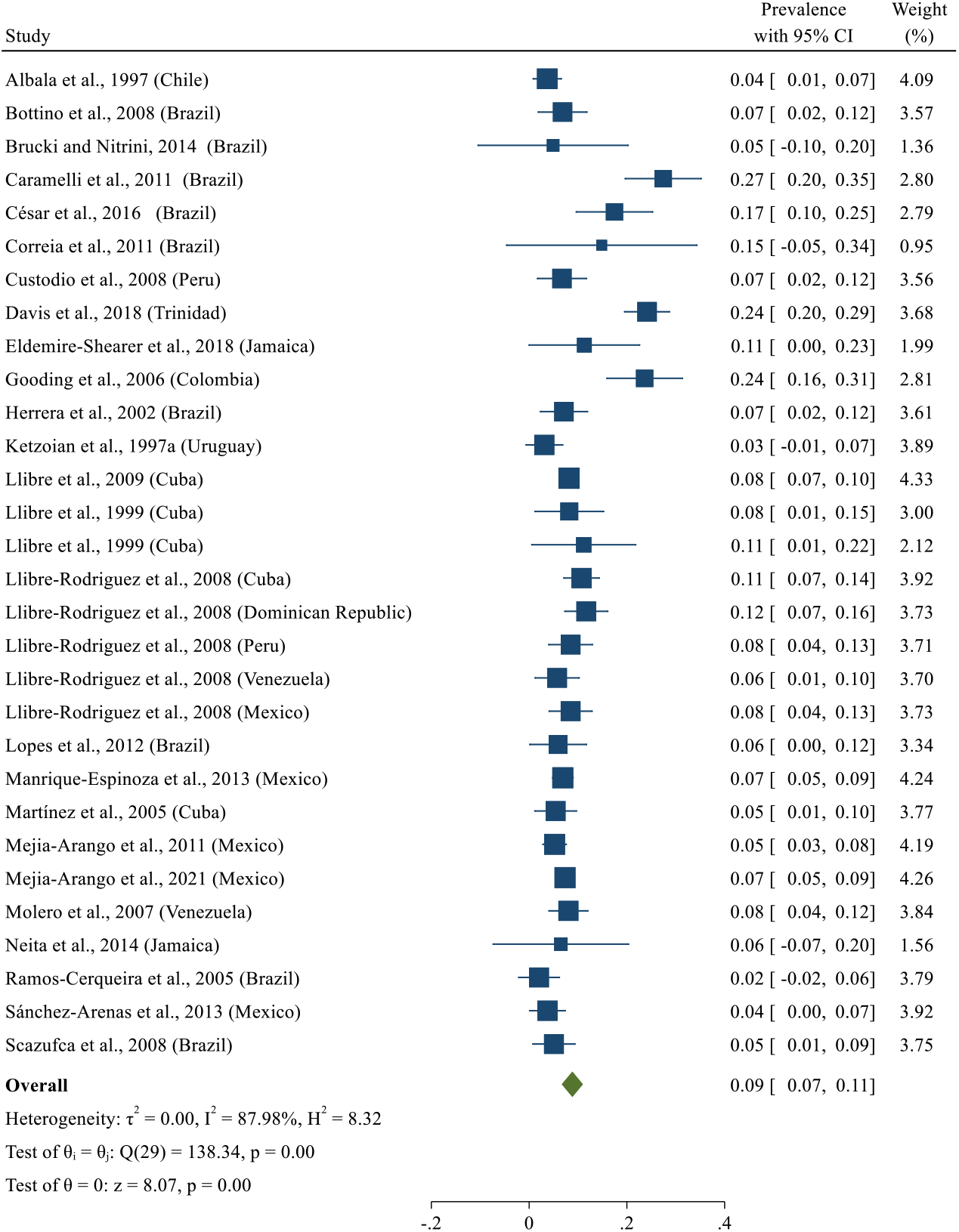
All-cause dementia prevalence after performing the leave-one-out method.

In the next step, we performed four sub-group meta-analyses stratifying by sex and including only those studies with enough information (n = 24 for the set of analyses of dementia prevalence to the overall participants and n = 23 for the set of analyses for each sex specifically for same-sex participants). We excluded Magalhães et al. (2008) as the leave-one-out from the previous section pointed out that this study considerably increased the overall estimate.

As displayed in Figure 5, the first analyses showed a pooled prevalence of dementia estimated of 3% (95% CI: 2% - 3%) for men and 5% (95% CI: 4% - 6%) for women, which was lower compared to the overall estimate which was based on all studies including those that did not report dementia prevalence by sex or that were flagged by the leave-one-out method.

**Figure 5.**
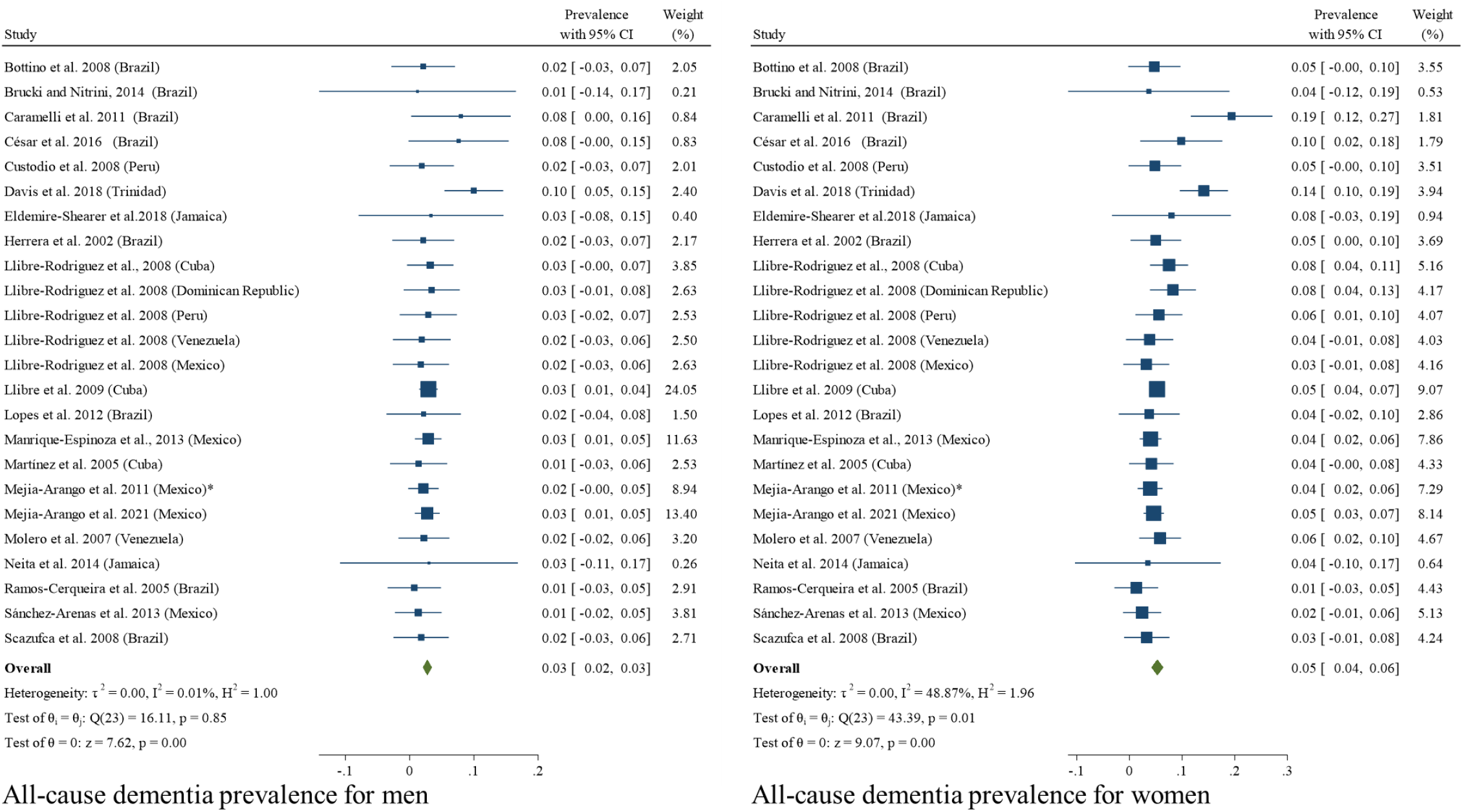
All-cause dementia prevalence by sex to the overall participants. *Note:*\* Additional information retrieved from Mejia-Arango et al. (2021).

The following sub-group analyses, similarly to previous ones, showed a higher prevalence for women (9%, 95% CI: 7% - 15%) than men (5%, 95% CI: 4% - 6%) when pooling prevalence of dementia by sex (see Figure 6).

**Figure 6.**
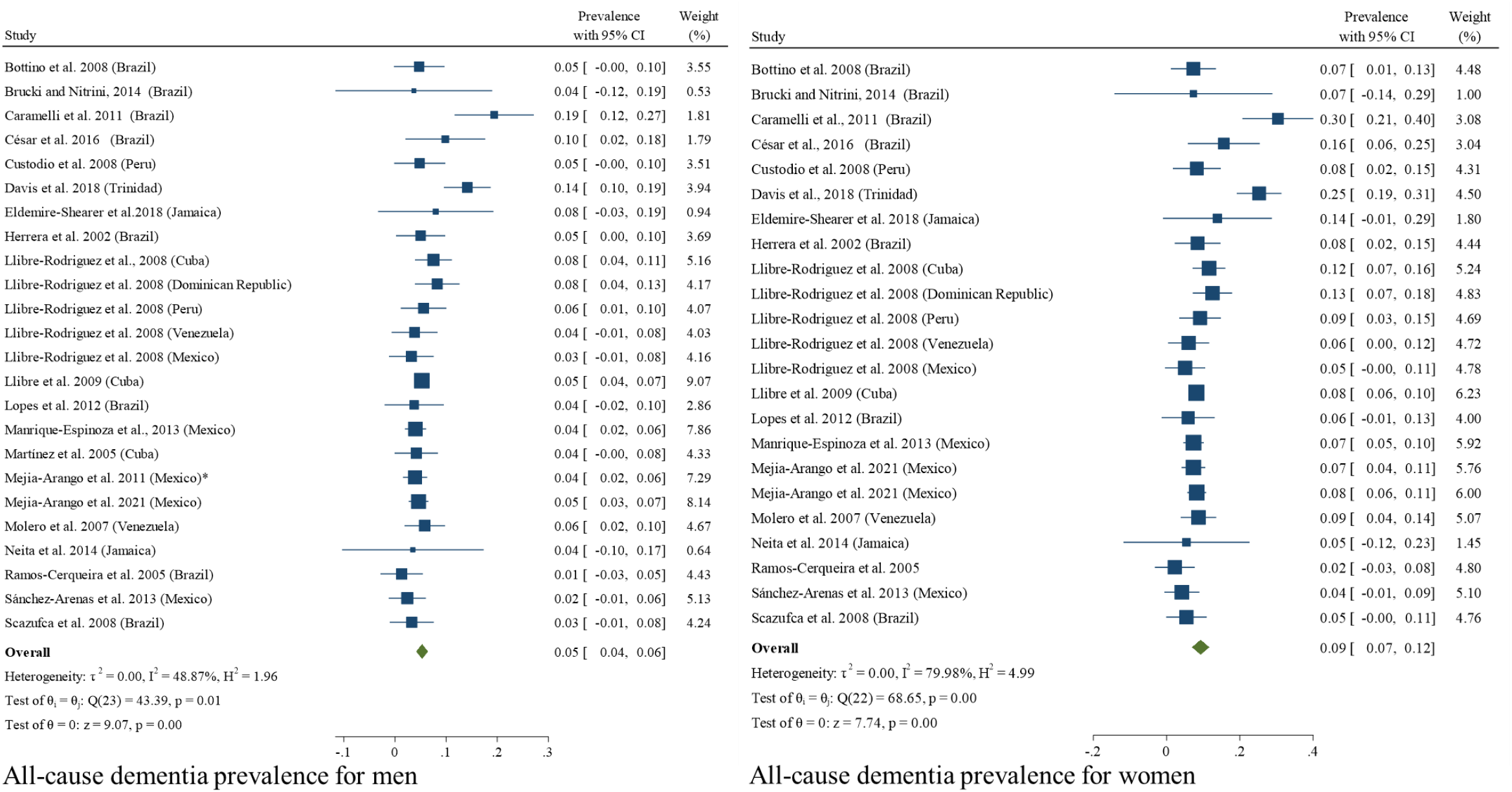
All-cause dementia prevalence by sex to the same-sex participants. *Note:*\* Additional information retrieved from Mejia-Arango et al. (2021).

Furthermore, we carried out two separated subgroup meta-analyses for rural and urban samples. Eight studies included rural samples, while 26 studies were based on urban samples. One of the articles previously excluded due to a fully rural sample, Magalhães et al. (2008), was included in the analysis to estimate the pooled prevalence in the rural area. As displayed in Figures 5 and 6, the pooled prevalence of all-cause dementia was 12% for rural areas (95% CI: 2% - 12%) and 8% for urban areas (95% CI: 6% - 10%). However, this result must be sanalysed with caution since only a small number of studies (n = 8) have been conducted among rural populations (see Figure 7-8).

**Figure 7.**
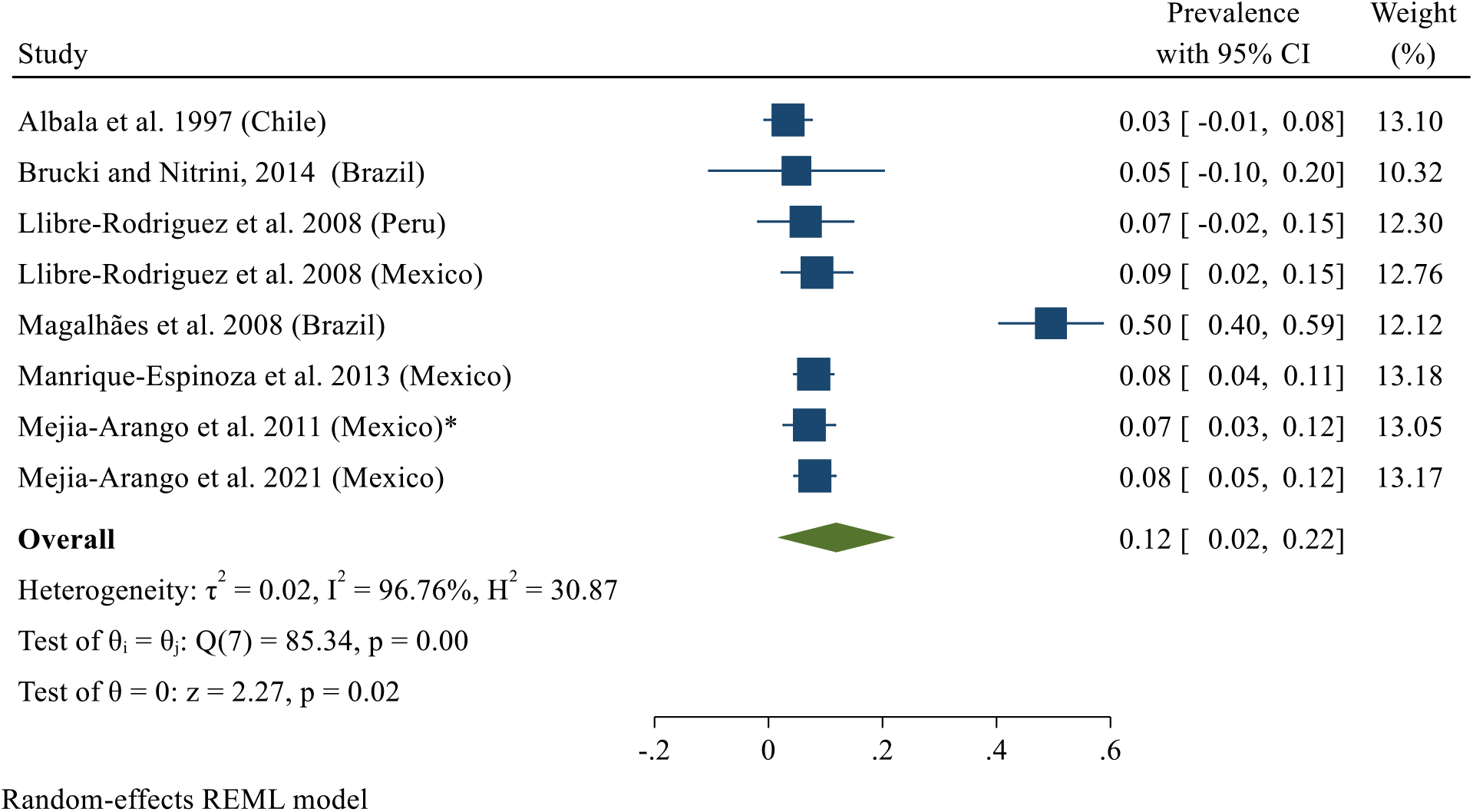
All-cause dementia prevalence by rural area. *Note:*\* Additional information retrieved from Mejia-Arango et al. (2021).

**Figure 8.**
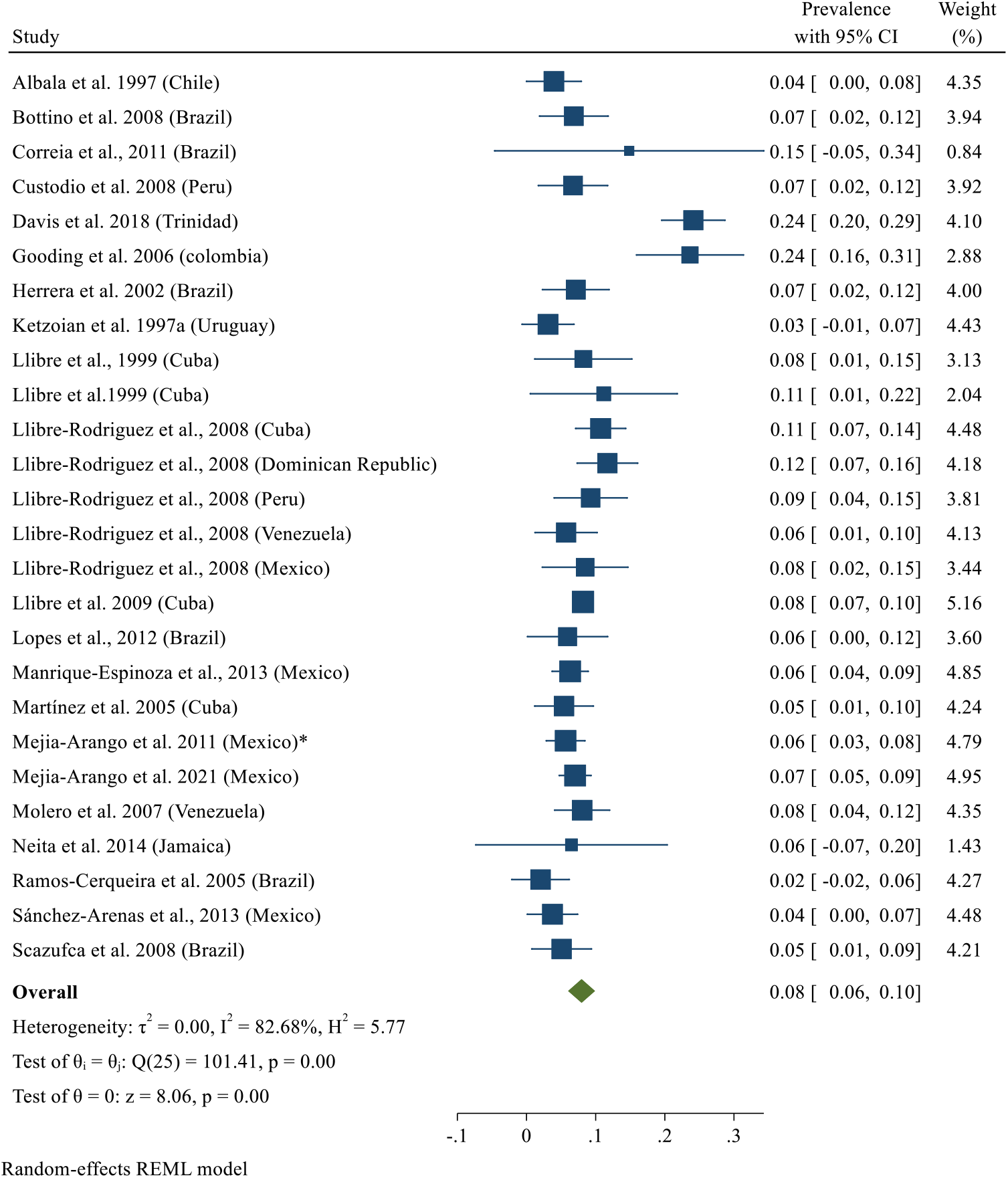
All-cause dementia prevalence by urban area. *Note:*\* Additional information retrieved from Mejia-Arango et al. (2021).

Finally, we performed two separated subgroups meta-analyses comprising those studies with complete data for educational level to explore the dementia prevalence for no formal education and at least one year of formal education. As displayed in Figures 9-10, our analyses revealed a prevalence of 22% (95% CI: 14% - 29%) for those participants without formal education and a prevalence of 10% (95% CI: 7% - 14%) for those with at least one year of formal education.

**Figure 9.**
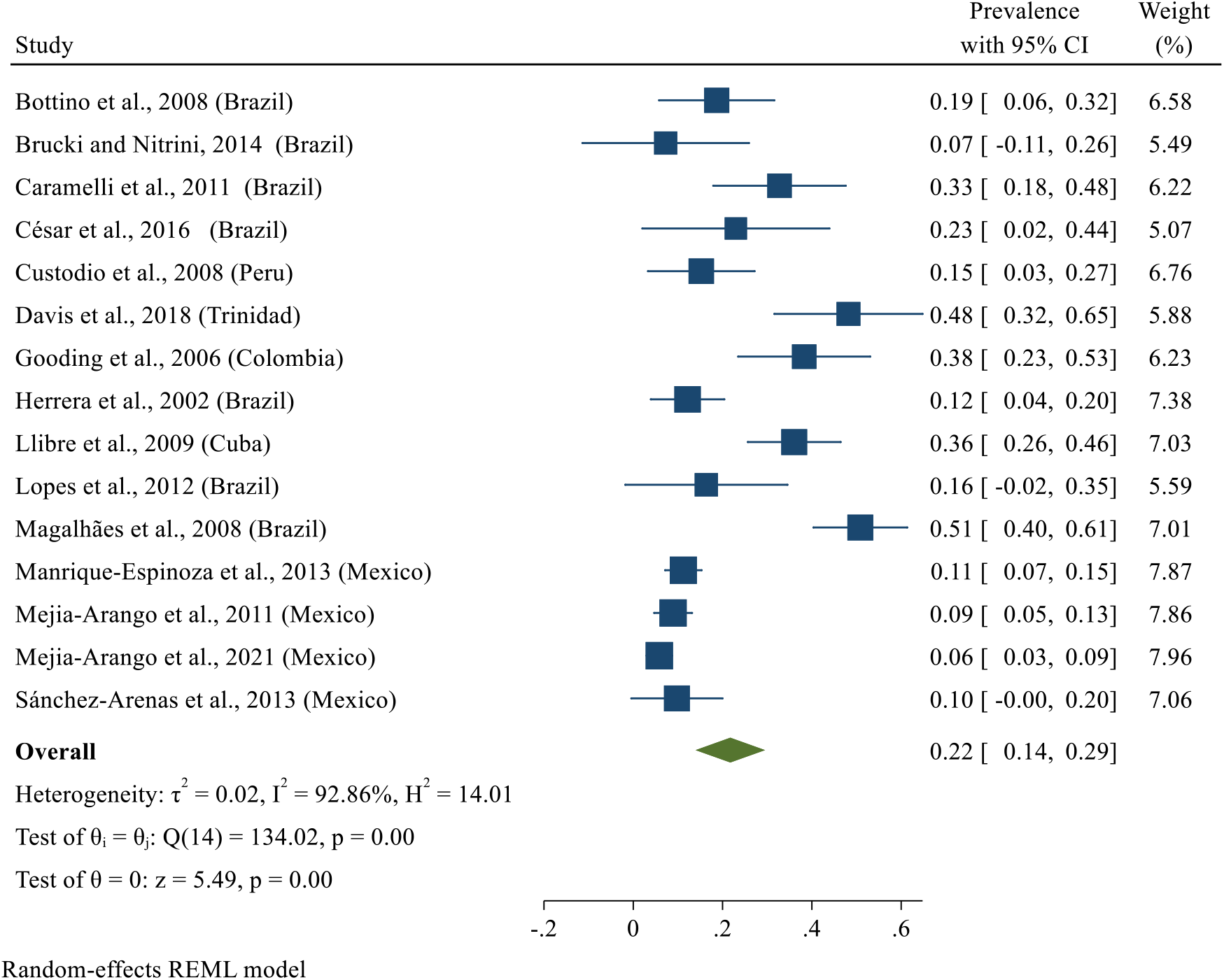
All-cause dementia prevalence for participants with no formal education.

**Figure 10.**
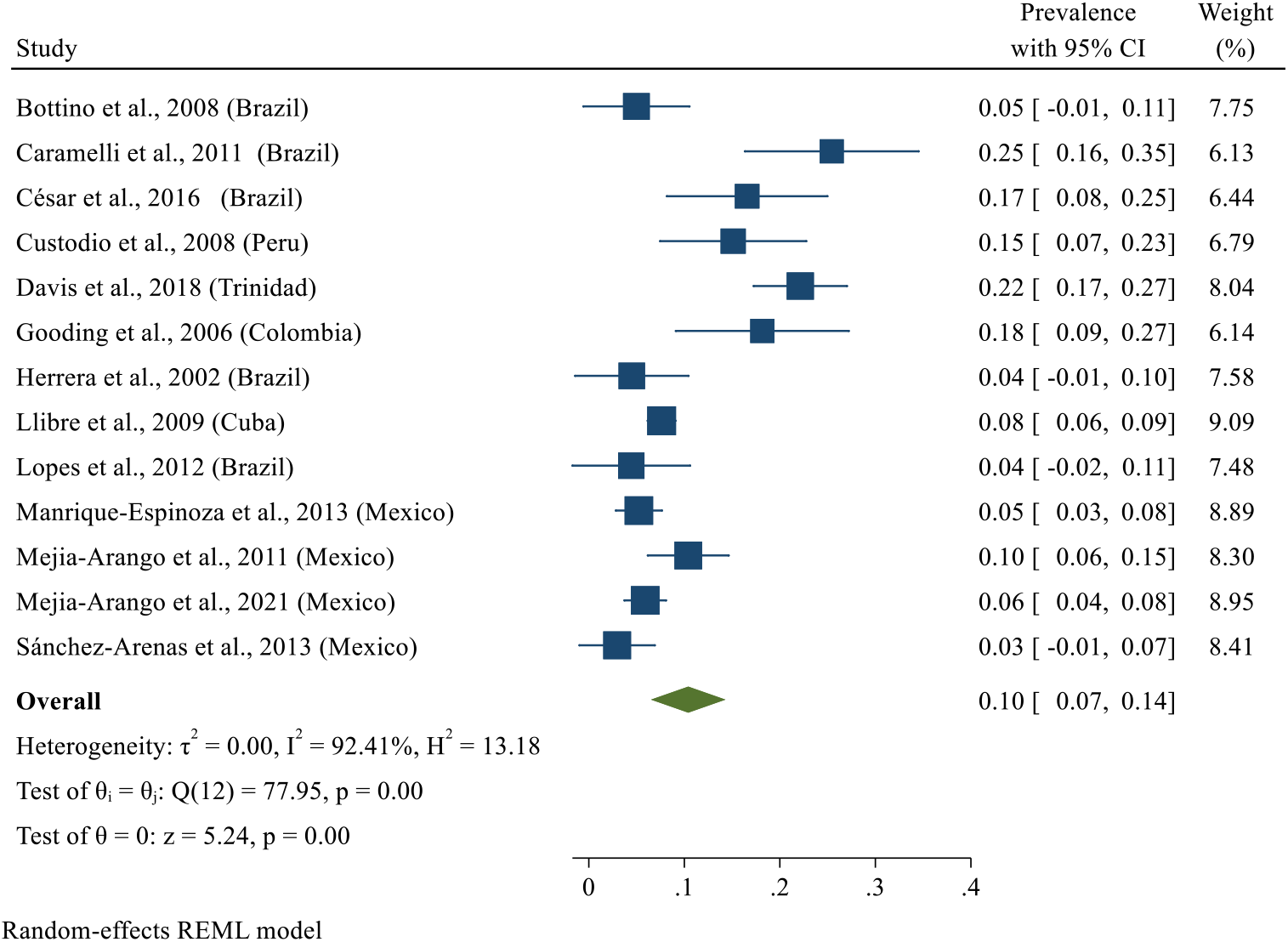
All-cause dementia prevalence for participants with at least one year of formal education.

#### Publication bias

Eggers’s test results were not significant for neither the pooled prevalence of all-cause dementia after performing the leave-one-out method (t = 1.88, *p* = 0.06), nor sex-specific prevalence of all-cause dementia to overall participants (women: *t* = 1.53, *p* = 0.12; men: *t* = 0.45, *p* = .66), to same-sex participants (women: *t* = 1.26, *p* = 0.21; men: *t* = 1.18, *p* = .24), for areas (rural: *t* = -0.56, *p* = 0.57; urban: *t* = 1.45, *p* = 0.15), or educational level (no formal education: *t* = 1.61, *p* = 0.11; at least one year of formal education: *t* = 2.01, *p* = 0.06), suggesting absence of publication bias. The corresponding funnel plots are provided in supplementary Figures S5-S13.

##### 3.5. Qualitative findings

We qualitatively verified that dementia prevalence was slightly higher in studies with more recent data collection, in the years between 2005 and 2015, compared to studies with data collection earlier than 2005 (Bottino et al., 2008; Brucki & Nitrini, 2014; Herrera et al., 2002; Ketzoian et al., 1997a; Llibre et al., 2009; Llibre et al., 1999; Lopes et al., 2012; Mejia-Arango & Gutierrez, 2011; Molero et al., 2007; Ramos-Cerqueira et al., 2005). Estimates of all-cause dementia prevalence by year of data collection are represented in Figure 11.

**Figure 11.**
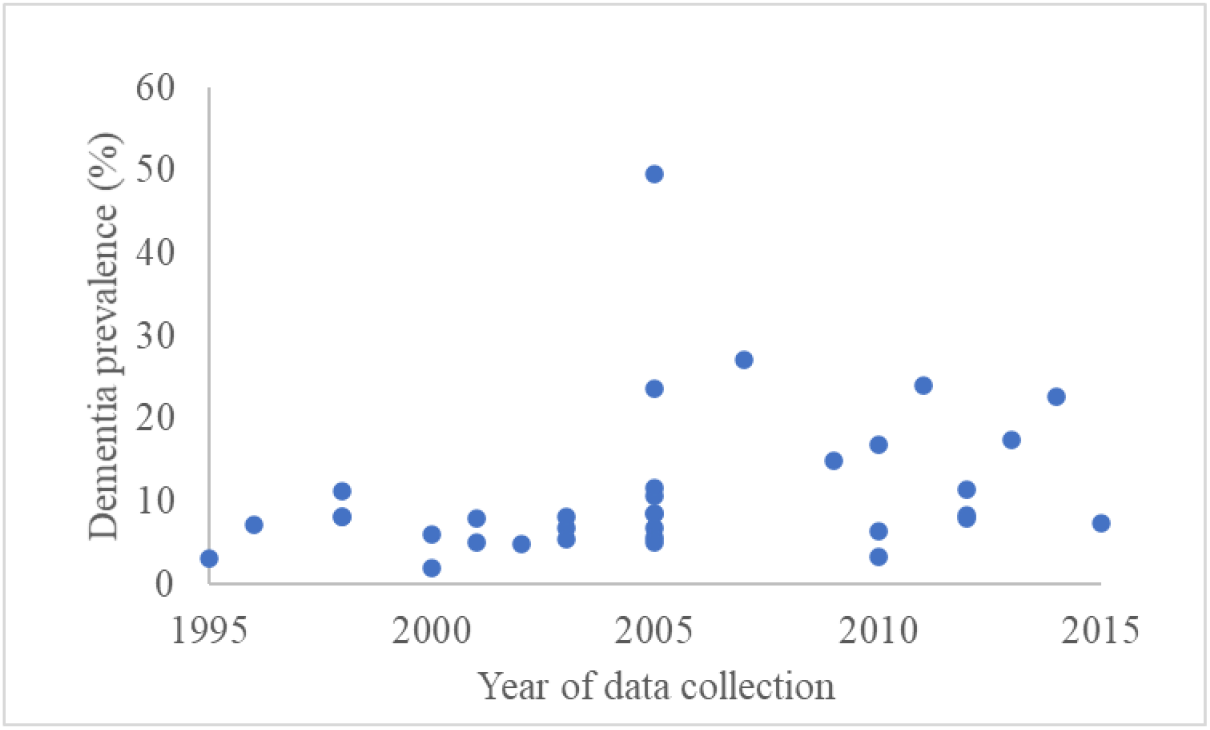
Dementia prevalence estimates across years of data collection.

Studies had slight variations regarding the age inclusion criteria. Two studies included participants aged 50+ years (Brucki & Nitrini, 2014; Pedraza et al., 2017), one article included participants aged 55+ years (Molero et al., 2017). In contrast, 13 studies included participants aged 60+ years (Bartoloni et al. 2014; Bottino et al. 2008; César et al. 2016; Gooding et al. 2006; Llibre et al., 1999; Lopes et al., 2012; Magalhães et al. 2008; Mejia-Arango et al. 2011; Neita et al. 2014; Vargas-Alarcón et al. 2016), eight included participants aged 65+ years (Correia et al. 2011; Custodio et al. 2008; Herrera et al. 2002; Llibre et al. 2009; Llibre-Rodriguez et al., 2008; Ramos-Cerqueira et al. 2005; Scazufca et al. 2008; Martínez et al. 2005), one included participants aged 70+ years (Davis et al., 2018) and one study included participants aged 75+ years (Caramelli et al., 2011). We thus report prevalence estimates by age group (50-59, 60-64, 65-69, 70-74, 75-79, 80+) in Table 2. Some studies reported a slightly higher prevalence of dementia at earlier ages (> 64), going from 5.3 up to 44% (César et al., 2016; Gooding et al., 2006; Magalhães et al., 2008; Pedraza et al., 2017; Vargas-Alarcón et al., 2016) in comparison with the remaining studies, in which the percentage was around 2-3% at this age range. In Pedraza et al. (2017), the study was designed to include participants with 60+ years old; however, some 50-59 years-old adults were non-randomly selected into the study, resulting in a potentially biased estimate of dementia prevalence at ages 50 to 59.

**Table 2.**
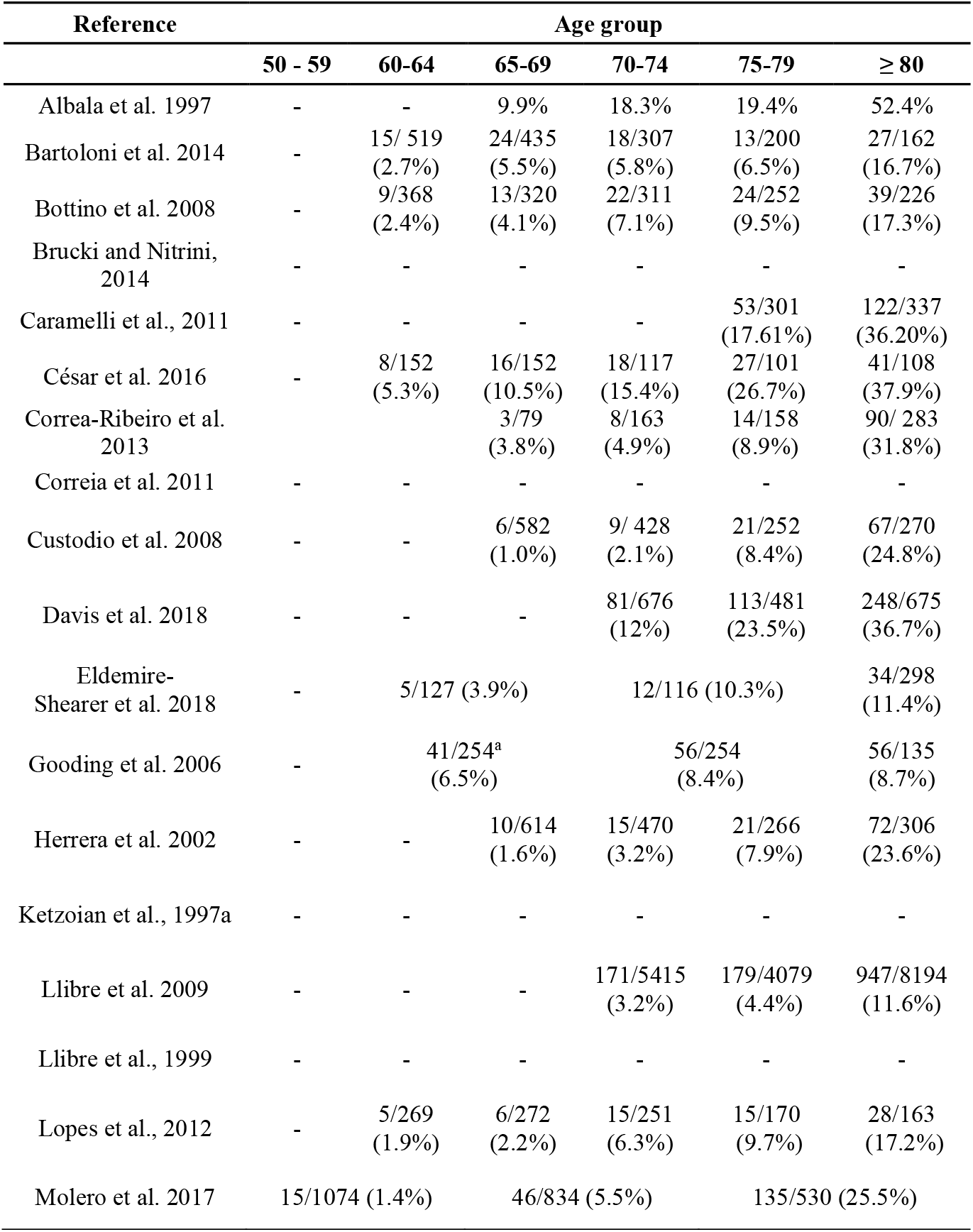
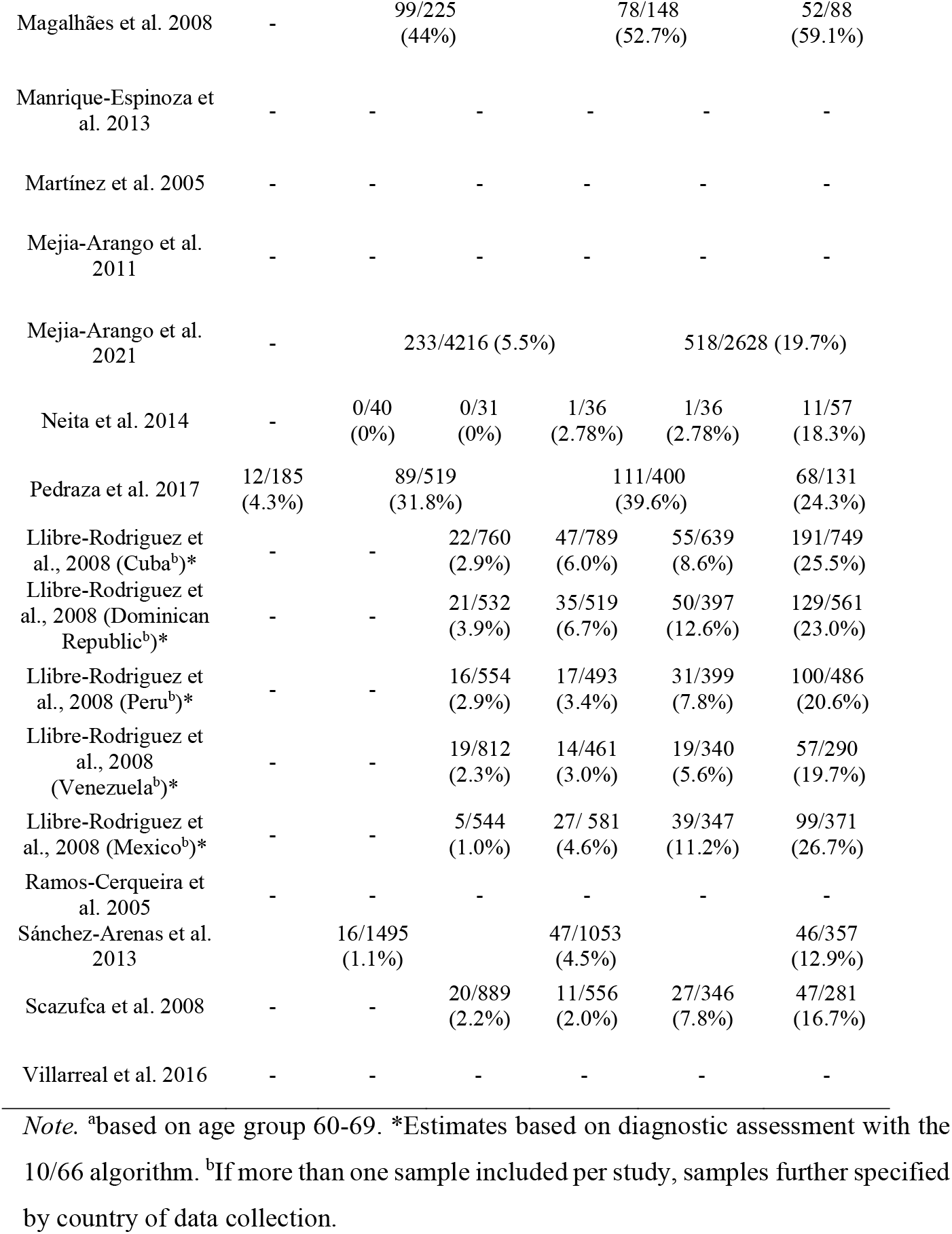
The number of study participants diagnosed with dementia against the total number of participants by age groups across the included studies.

## Discussion

This systematic review and meta-analysis of the prevalence of all-cause dementia in LAC countries used strict inclusion criteria for dementia diagnosis and included research published in the predominant languages spoken in this region, resulting in a comprehensive evidence base and providing updated and robust estimates of dementia prevalence for a region inhabited by around 8.4% of the world population. In our main analysis, we observed a prevalence of all-cause dementia of 11% for adults aged 50 or more years old. However, when excluding those articles detected as influencing the results by the leave-one-out method, the new prevalence estimate was 9%, still higher compared with estimates in high-income countries, for instance, USA with 8.2% with participants aged 65+ years (Koller & Bynum, 2015) or Europe with 7.1% (Bacigalupo et al., 2018) including participants with ages 55 years old and above.

Lower levels of education might explain the higher prevalence of dementia among older adults of LAC countries (Cao et al., 2020; Custodio et al., 2017; Mukadam et al., 2019), reflected in the higher number of illiteracy and low educational attainment across the selected studies, added to low socioeconomic levels, and limited access to primary health care and poor control of cardiovascular risk factors. In fact, the pooled analysis based on studies with rural samples showed a dementia prevalence of 14%, compared to 9% based on studies with urban samples. Moreover, not only the level of education but also lower-skilled occupations which are predominantly manual, in the case of older adults in rural areas, are related to a higher prevalence of dementia (Alvares Pereira et al., 2021; Hyun et al., 2020). However, it is important to mention that more studies with rural samples are necessary since only seven included rural populations.

We observed a high heterogeneity between studies, possibly leading to the high variation in the crude prevalence, ranging from 2.02% to 49.57%. This disparity could be explained by several factors, such as samples coming from rural or urban regions, different socioeconomic backgrounds, ages, and also by the methods of diagnosing dementia, as the instruments and assessment strategies still broadly varied among studies – despite the rigorous inclusion criteria applied in this study. To arrive at more robust findings, it would be desirable to plan cross-national analyses through a consortium spanning different LAC countries and apply the same study protocol in order to reduce heterogeneity across studies.

In a subsample of studies with sex-stratified estimates of dementia prevalence, we verified that dementia prevalence rate was higher among women than men. These findings seem to be explained by differences in higher life expectancy in women compared to men (Carter et al., 2012; Parra et al., 2018). Adding to this explanation, one should not minimise gender inequalities in education, work, and health, especially in low- and middle-income countries, meaning lower access to education, lower-income jobs, and lower access to health services by women (Carter et al., 2012; Snyder et al., 2016; Zurique Sánchez et al., 2019).

Given that high-income countries present a decrease in number of dementia prevalence lately (Langa et al., 2017; Matthews et al., 2013; Satizabal et al., 2016), we qualitatively investigated secular trends in LAC. Contrarily, our analysis showed an increase in dementia prevalence estimates in the studies with more recent data collection, which could be explained by increases in life expectancy in LAC, as there has been an increase of approximately seven years in life expectancy between 1990 to 2020 (United Nations, 2019). Additionally, increases in unhealthy lifestyles and prevalence of chronic conditions may be driving the increases in dementia prevalence estimates in more recent studies. For example, a worse diet is associated with higher levels of cardiovascular risk factors leading to neurodegenerative damage (Feigin et al., 2016). Further, factors related to an unhealthy lifestyle, i.e., high body mass index, high fasting plasma glucose, smoking, and high intake of sugar-sweetened beverages, are strongly linked to Alzheimer’s disease and other dementias (Global Burden Disease Study, 2019). Consistent with secular increases in dementia prevalence identified in this meta-analysis, Ribeiro et al. (in press) showed that in one of the LAC countries, in Brazil, respondents aged 60 or more years old were more likely to report diabetes, hypertension, and overweight/obesity in 2015 compared to 2000-2010.

Regarding the educational level of participants of the studies included in this review, we quantitatively observed that older adults with no formal education presented a much higher prevalence of dementia (23 % *vs*. 10%) in comparison with older adults with at least one year of formal education, suggesting the protective effect of education against dementia. However, it was not possible to include all the studies in the pooled prevalence by education due to the lack of information provided in some studies. Nevertheless, it was possible to qualitatively observe that the samples were predominantly composed of adult illiterates or up to 4 years of schooling, reflecting the reality of older people living in many LAC countries and may underlie the high dementia prevalence found in this review. In fact, previous studies exploring the contribution of educational level to dementia suggested inverse associations between the number of years of education and the risk of developing dementia (Custodio et al., 2008; Langa et al., 2017; Nitrini et al., 2009). In line with this, several studies propose that education can promote an increase in cognitive reserve, i.e., increased brain adaptability to compensate neuropathological and vascular damage before dementia symptoms appear (Mondini et al., 2016; Prince et al., 2012).

Regarding the methodological quality of studies assessed using the JBI critical appraisal checklist for studies reporting prevalence data, most studies generally complied well with the different aspects of the checklist. However, some studies lacked information on recruitment of participants, standardisation of measurements or response rate. Additionally, some studies did not report confidence intervals when describing statistical results.

### Limitations and strengths of this study

This study provides updated and stratified estimates of dementia prevalence, firstly, for the LAC countries as a world region with less evidence on the epidemiology of dementia compared to other regions, such as Europe and the United States. Further, synthesising evidence from multiple studies that results in the most comprehensive review of dementia prevalence in LAC countries to date, our findings provide some confidence on the robustness of the dementia prevalence estimates. In fact, dementia is a health condition for which data scarcity and limited data quality are hampering the provision of serious estimates worldwide (Launer, 2019). Another strength of this study is the inclusion of studies with clear diagnostic criteria for dementia, as well as studies published in all languages predominantly spoken in LAC countries. The high and rising prevalence of dementia in LAC showed the importance of recognising dementia as a growing public health priority, especially considering that LAC countries have limited resources to deal with this burden. For this reason, it is crucial to develop well-structured action plans for dementia in this region (Parra et al., 2018, 2021; Zurique Sánchez et al., 2019). To put this into practice, it would require early and broad access to dementia diagnosis with practices based on scientific consensus, as well as training of multidisciplinary teams.

One limitation of this systematic review was the heterogeneity of the included studies, in addition to the small number of investigations on dementia prevalence published in LAC countries. Actually, we did not find studies performed in the following countries: Guatemala, Belize, El Salvador, Nicaragua, Costa Rica, Panama, Antigua and Barbuda, The Bahamas, Barbados, Dominica, Grenada, Haiti, Kitts and Nevis, Lucia, Vincent and the Grenadines, Ecuador, Bolivia, Paraguay, Guyana, and Suriname. Filling these gaps in evidence for a significant part of this world region could be included in the scientific research agenda of large public health funders. Specifically, future studies could consider following strict criteria for diagnosis of dementia, such as DSM or ICD criteria, having more than one phase to screen and assess participants, performing different analyses considering age, sex, education, socioeconomic status, and assess possible comorbidities, such as cardiovascular risk factors.

### Conclusions

Providing robust and updated dementia prevalence estimates for the large world region of LAC countries, this systematic review shows that dementia prevalence differs in the LAC countries according to sex and area, similar to evidence from other world regions such as Europe and the United States, although the prevalence rate is higher in LAC. Our meta-analysis point to possible increases in dementia prevalence in LAC studies with more recent data collection, questioning if the secular decreases in dementia incidence and prevalence observed in high-income countries hold in all world regions and are a strong reminder of the public health action on dementia in LAC countries. Considering rapidly ageing populations in the LAC countries, providing more robust estimates for the countries missing in this review would help estimate the societal and economic burden of dementia more accurately and assist in public health planning.

## Data Availability

All data produced in the present study are available upon reasonable request to the authors

