## Supplementary Material for "Systematic review and meta-analyses on the prevalence of dementia in Latin America and Caribbean countries: Exploring sex, rurality, age, and education as possible determinants"

The search terms for each database and number of found articles

**Updated search on 13 – 08– 2021**

**Pubmed (n = 961)**

(dementia [MeSH]) AND (Prevalence OR Epidemiology) AND ("Latin America" OR “South America” OR Caribbean OR Argentina OR Bolivia OR Brazil OR Chile OR Colombia OR "Costa Rica" OR Cuba OR Ecuador OR "El Salvador" OR Guatemala OR Haiti OR Honduras OR Mexico OR Nicaragua OR Panama OR Paraguay OR Peru OR "Dominican Republic" OR Uruguay OR Venezuela OR Jamaica OR “Trinidad and Tobago” OR Guyana OR Suriname OR Belize OR Bahamas OR Barbados OR “Saint Lucia” OR Grenada OR “St. Vincent and Grenadines” OR “Antigua and Barbuda” OR Dominica OR “Saint Kitts and Nevis”)

**Web of knowledge (n = 528)**

(Dementia) AND (Prevalence OR Epidemiology) AND ("Latin America" OR “South America” OR Caribbean OR Argentina OR Bolivia OR Brazil OR Chile OR Colombia OR "Costa Rica" OR Cuba OR Ecuador OR "El Salvador" OR Guatemala OR Haiti OR Honduras OR Mexico OR Nicaragua OR Panama OR Paraguay OR Peru OR "Dominican Republic" OR Uruguay OR Venezuela OR Jamaica OR “Trinidad and Tobago” OR Guyana or Suriname OR Belize OR Bahamas OR Barbados OR “Saint Lucia” OR Grenada OR “St. Vincent and Grenadines” OR “Antigua and Barbuda” OR Dominica OR “Saint Kitts and Nevis”)

**Scopus (n = 538)**

TITLE-ABS-KEY ( ( ( dementia ) AND ( prevalence OR epidemiology ) AND ( "Latin America" OR "South America" OR caribbean OR argentina OR bolivia OR brazil OR chile OR colombia OR "Costa Rica" OR cuba OR ecuador OR "El Salvador" OR guatemala OR haiti OR honduras OR mexico OR nicaragua OR panama OR paraguay OR peru OR "Dominican Republic" OR uruguay OR venezuela OR jamaica OR "Trinidad and Tobago" OR guyana OR suriname OR belize OR bahamas OR barbados OR "Saint Lucia" OR grenada OR "St. Vincent and Grenadines" OR "Antigua and Barbuda" OR dominica OR "Saint Kitts and Nevis" ) ) )

**Lilacs - (n= 1290)**

mh:((dementia) OR (demencia) OR (demência) AND (prevalenc*) OR (prevalência) OR (epidemiolog*)) AND ( db:("LILACS"))

**SciELO (n= 79)**

( dementia ) AND ( prevalence OR epidemiology ) AND ( "Latin America" OR "South America" OR caribbean OR argentina OR bolivia OR brazil OR chile OR colombia OR "Costa Rica" OR cuba OR ecuador OR "El Salvador" OR guatemala OR haiti OR honduras OR mexico OR nicaragua OR panama OR paraguay OR peru OR "Dominican Republic" OR uruguay OR venezuela OR jamaica OR "Trinidad and Tobago" OR guyana OR suriname OR belize OR bahamas OR barbados OR "Saint Lucia" OR grenada OR "St. Vincent and Grenadines" OR "Antigua and Barbuda" OR dominica OR "Saint Kitts and Nevis" )

**Table S1.** JBI critical appraisal checklist assessment results.

| Study Author and Year | Questions | | | | | | | | | |
|---|---|---|---|---|---|---|---|---|---|---|
| | Was the sample representative? | Were participants appropriately recruited? | Was the sample size adequate? | Are the subjects and setting described in detail? | Data analysis conducted with sufficient sample coverage? | Have valid methods been used for dementia identification? | Was dementia measured in a standard way? | Was there appropriate statistical analysis? | Was the response rate adequate? | Score |
| Albala et al. 1997* | Yes | Yes | Yes | No | No | Yes | Unclear | No | Yes | 5 |
| Bartoloni et al. 2014 | No | Yes | Yes | Yes | Yes | Yes | Yes | No | Yes | 7 |
| Bottino et al. 2008 | Yes | Yes | Yes | Yes | Yes | Yes | Yes | Yes | Yes | 9 |
| Brucki and Nitrini, 2014 | Yes | Unclear | Yes | Yes | Yes | Yes | Yes | No | Unclear | 5 |
| Caramelli et al., 2011 | Yes | Yes | Yes | Yes | Yes | Yes | Yes | No | Unclear | 7 |
| César et al. 2016[a] | Yes | Yes | Yes | Yes | Yes | Yes | Yes | Yes | Yes | 9 |
| Correa Ribeiro et al., 2013 | No | Yes | Yes | Yes | Yes | Yes | Yes | Yes | Yes | 7 |
| Correia et al. 2011 | Yes | Unclear | Yes | Yes | Yes | Yes | Yes | Yes | Yes | 8 |
| Custodio et al. 2008 | Yes | Yes | Yes | Yes | Yes | Yes | Yes | Yes | Yes | 9 |
| Davis et al. 2018 | Yes | Yes | Yes | Yes | Yes | Yes | Yes | Yes | Yes | 9 |
| Eldemire-Shearer et al. 2018 | Yes | Yes | Yes | No | Yes | Yes | Unclear | No | Yes | 6 |
| Gooding et al. 2006 | Yes | Yes | Yes | Yes | Yes | Yes | Yes | No | Unclear | 7 |
| Herrera et al. 2002 | Yes | Yes | Yes | Yes | Yes | Yes | Yes | No | Yes | 8 |
| Ketzoian et al. 1997a* | Yes | Yes | Yes | No | Yes | Yes | Yes | No | Unclear | 6 |

| | | | | | | | | | | |
|---|---|---|---|---|---|---|---|---|---|---|
| Llibre et al. 2009 | Yes | Yes | Yes | Yes | Yes | Yes | Yes | Yes | Yes | 9 |
| Llibre et al., 1999 | Yes | Yes | Yes | No | Yes | Yes | Yes | Yes | Unclear | 7 |
| Lopes et al., 2012 | Yes | Unclear | Yes | Yes | Yes | Yes | Yes | Yes | Yes | 8 |
| Macías Ortega et al. 2012 | Yes | Yes | Yes | Yes | Yes | Unclear | Unclear | No | Unclear | 5 |
| Molero et al., 2007 | Yes | Yes | Yes | Yes | Yes | Yes | Yes | Yes | Yes | 9 |
| Magalhães et al. 2008 | Yes | Yes | Yes | Yes | Yes | Yes | Unclear | Yes | Yes | 8 |
| Manrique-Espinoza et al., 2013 | Yes | Yes | Yes | Yes | Yes | Yes | Yes | No | Unclear | 7 |
| Martínez Querol et al. 2005 | Yes | Yes | Yes | Yes | Yes | Yes | Unclear | No | Unclear | 6 |
| Mejia-Arango and Gutierrez. 2011 | Yes | Yes | Yes | Yes | Yes | Yes | Unclear | Yes | Yes | 8 |
| Mejia-Arango et al. 2021 | Yes | Yes | Yes | Yes | Yes | Yes | Unclear | Yes | Yes | 8 |
| Neita et al. 2014 | Yes | Yes | Yes | No | Yes | Yes | Unclear | No | Unclear | 5 |
| Pedraza et al. 2017 | Unclear | Unclear | Yes | Yes | Yes | Yes | Unclear | Yes | Unclear | 5 |
| Llibre-Rodriguez et al., 2008 | Yes | Yes | Yes | Yes | Yes | Yes | Yes | Yes | Yes | 9 |
| Ramos-Cerqueira et al. 2005 | Yes | Unclear | Yes | No | No | Yes | Yes | No | Yes | 5 |
| Sánchez-Arenas et al., 2013 | Yes | Yes | Yes | Yes | Yes | Yes | Unclear | Yes | Unclear | 7 |
| Scazufca et al. 2008 | Yes | Yes | Yes | Yes | Yes | Yes | Yes | Yes | Yes | 9 |
| Velázquez-Brizuela et al. 2014 | Yes | Yes | Yes | Yes | Yes | No | Yes | Yes | Unclear | 7 |

| | | | | | | | | | |
|---|---|---|---|---|---|---|---|---|---|
| Villarreal et al., 2016 | No | Yes | Yes | Yes | Yes | Yes | Unclear | No | Unclear | 5 |

Note: a – Some information was retrieved from Cesar (2014) Doctoral thesis. *Information provided by the authors.

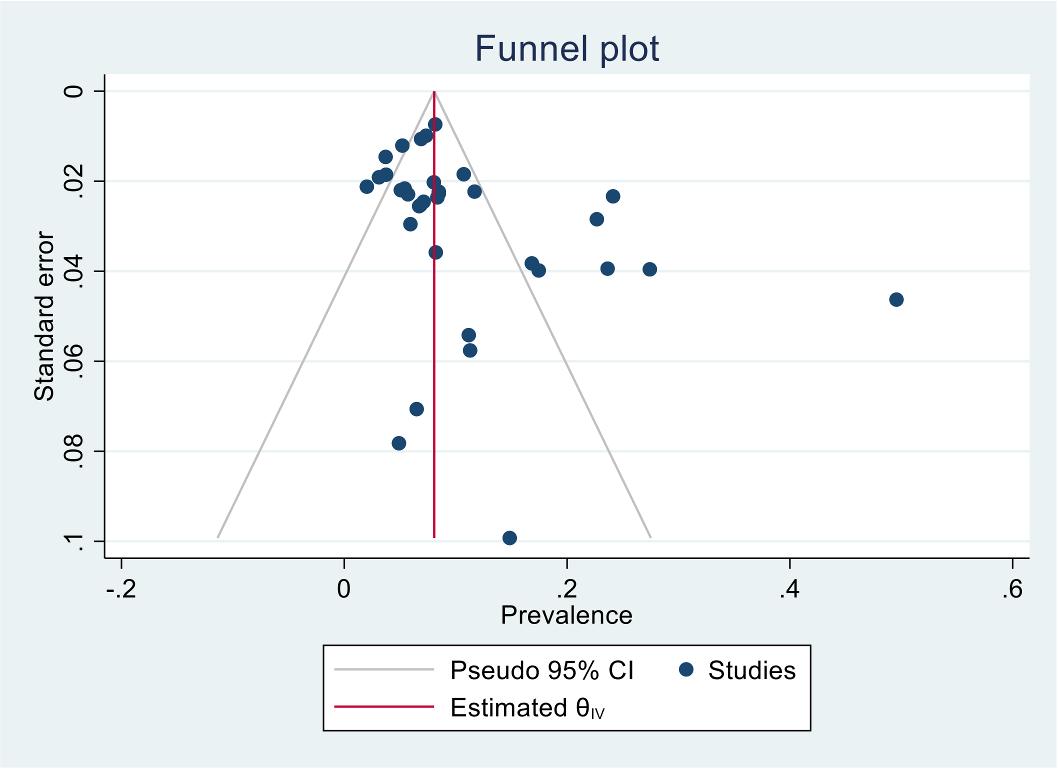


Figure S1: Funnel plot for meta-analysis of the prevalence of all-cause dementia including all selected studies

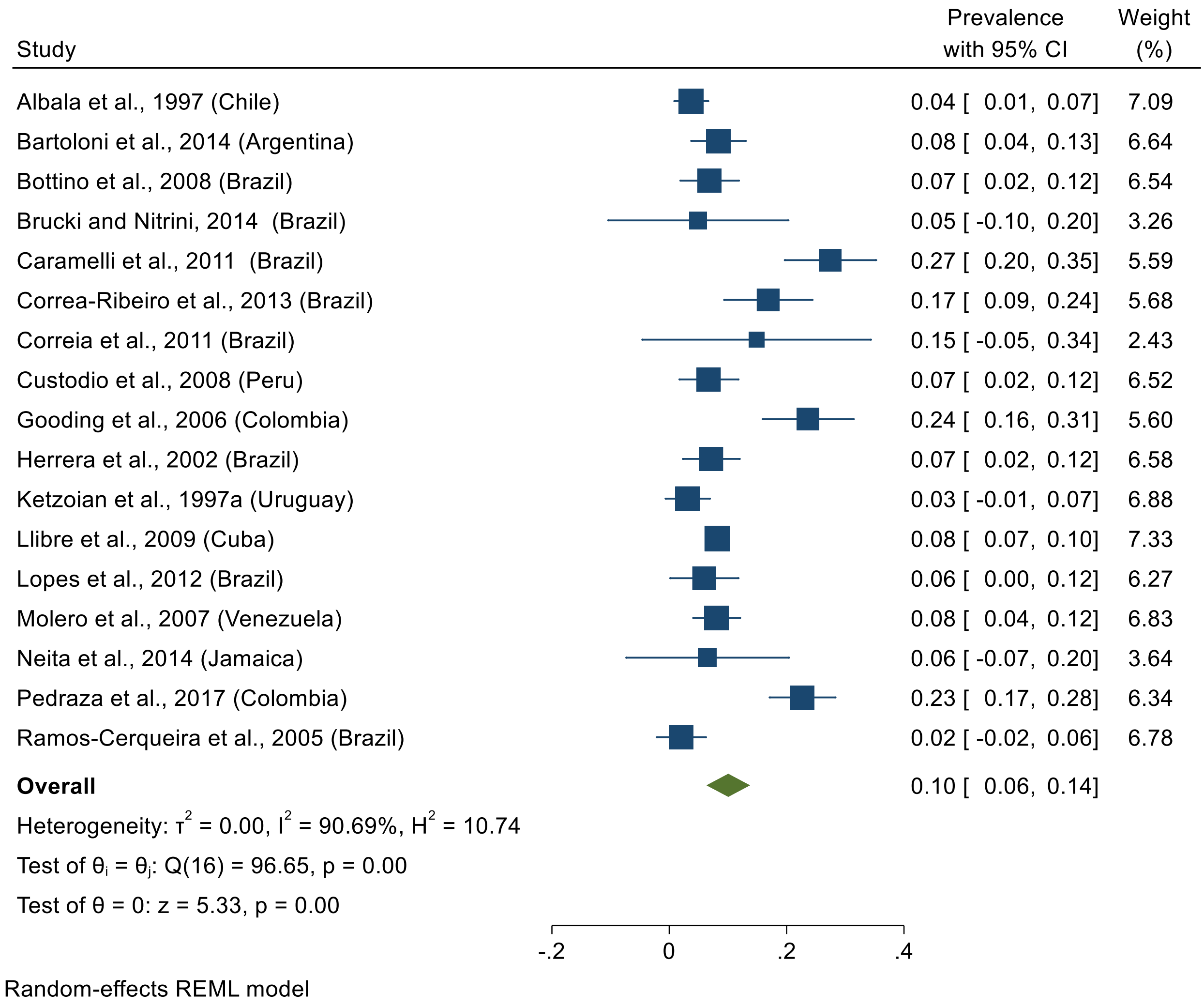


Figure S2. Meta-analysis of prevalence of all-cause dementia excluding studies with one-phase diagnosis.

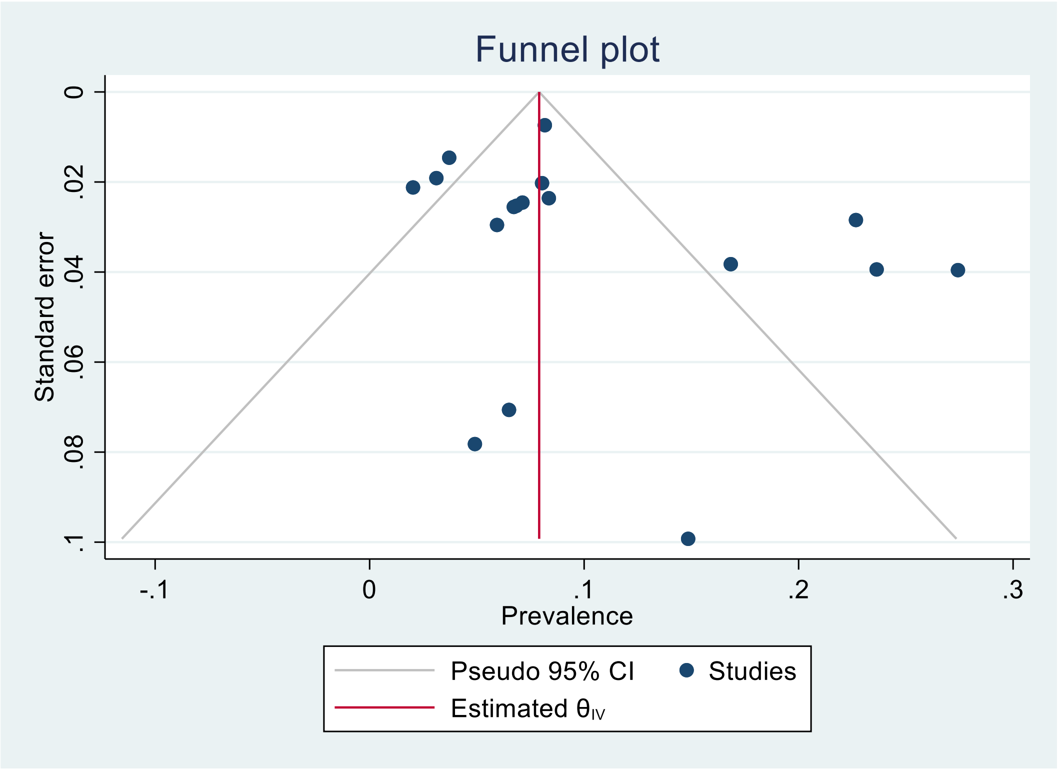


Figure S3: Funnel plot for meta-analysis of the prevalence of all-cause dementia excluding studies with one-phase diagnosis.

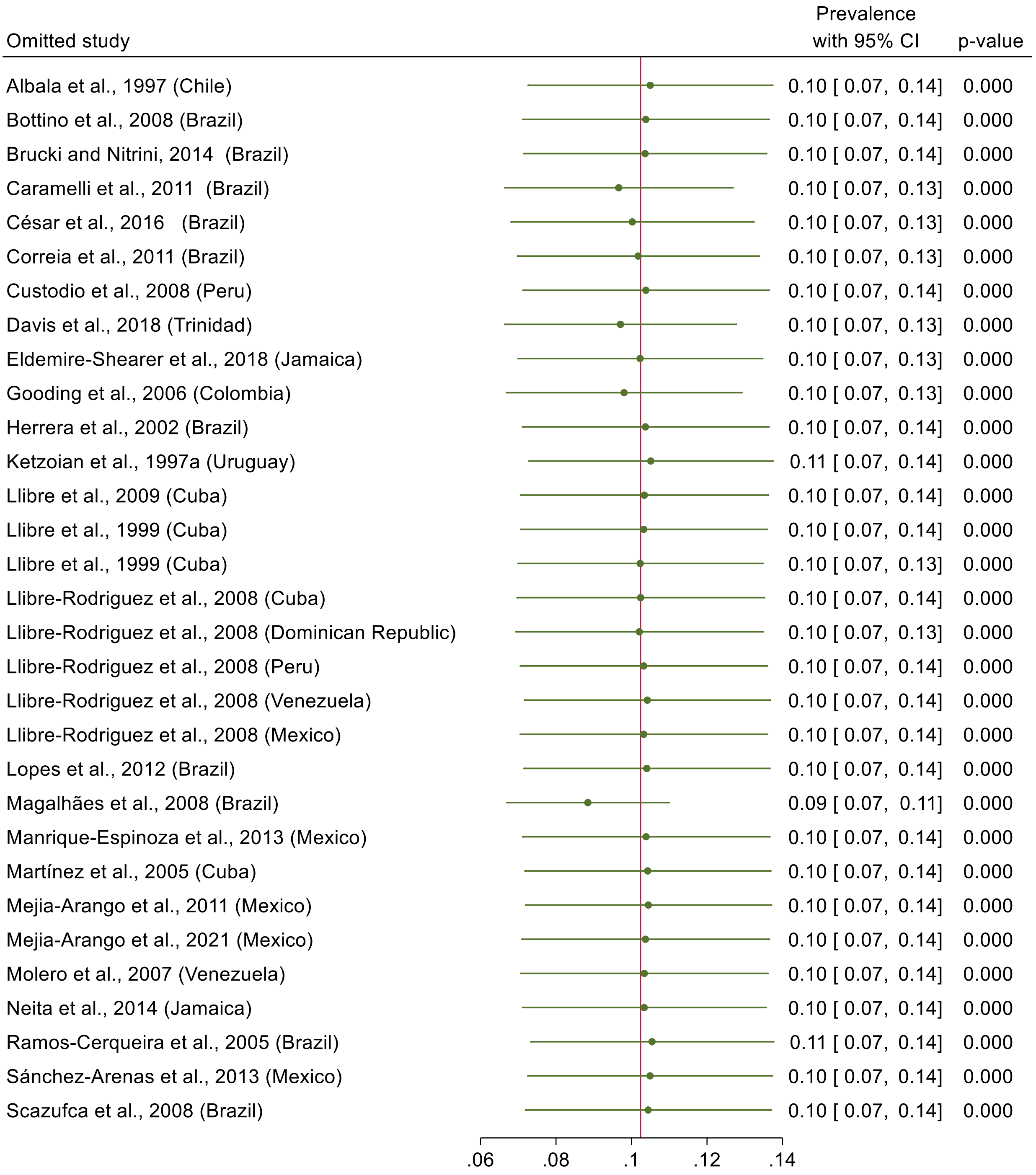


Figure S4. Leave-one-out method for all-cause dementia.

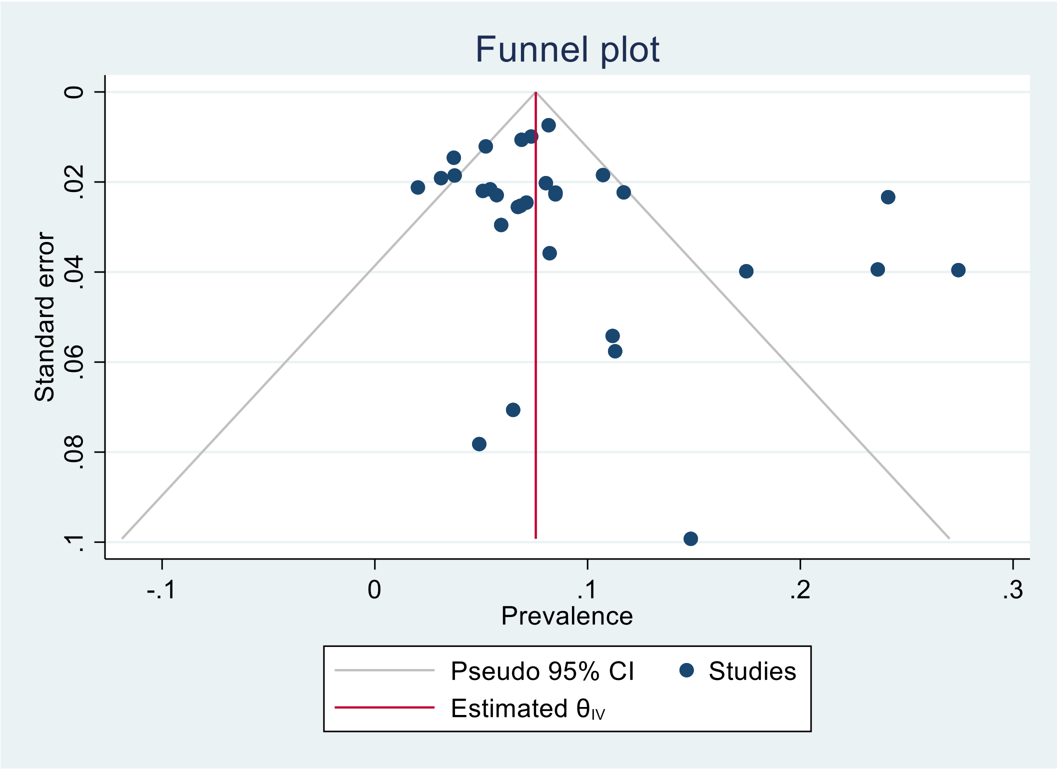


Figure S5: Funnel plot for meta-analysis of the prevalence of all-cause dementia after performing the leave-one-out method.

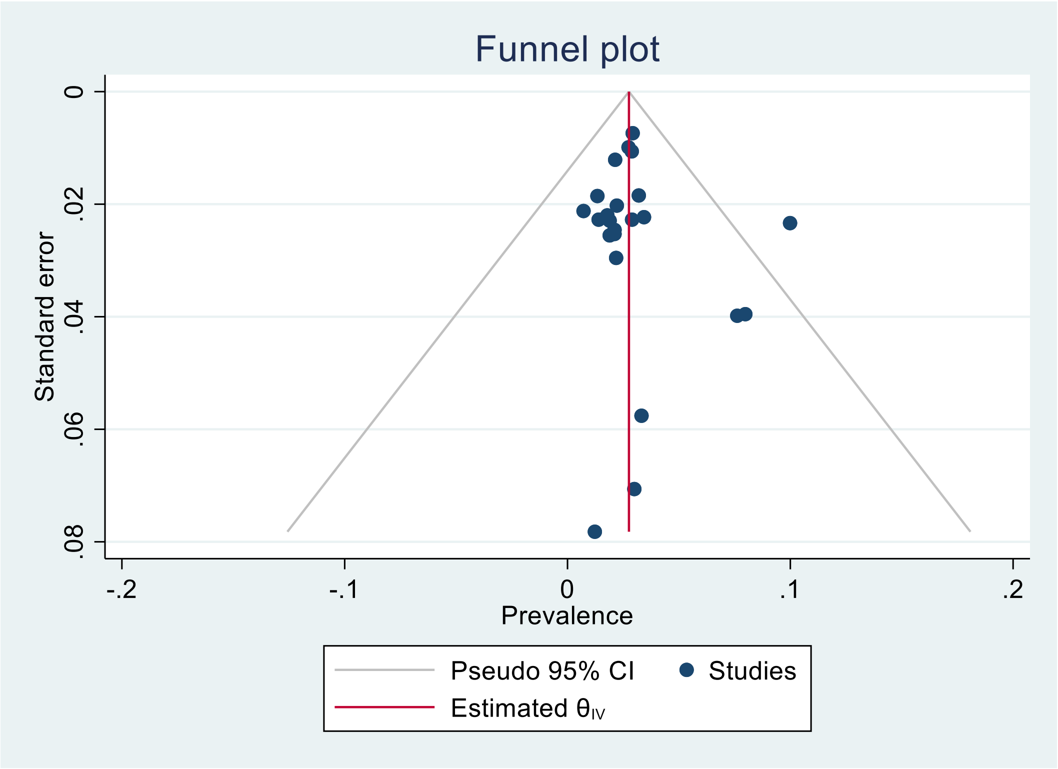

Figure S6: Funnel plot for meta-analysis of the prevalence of all-type dementia for men for the overall participants.

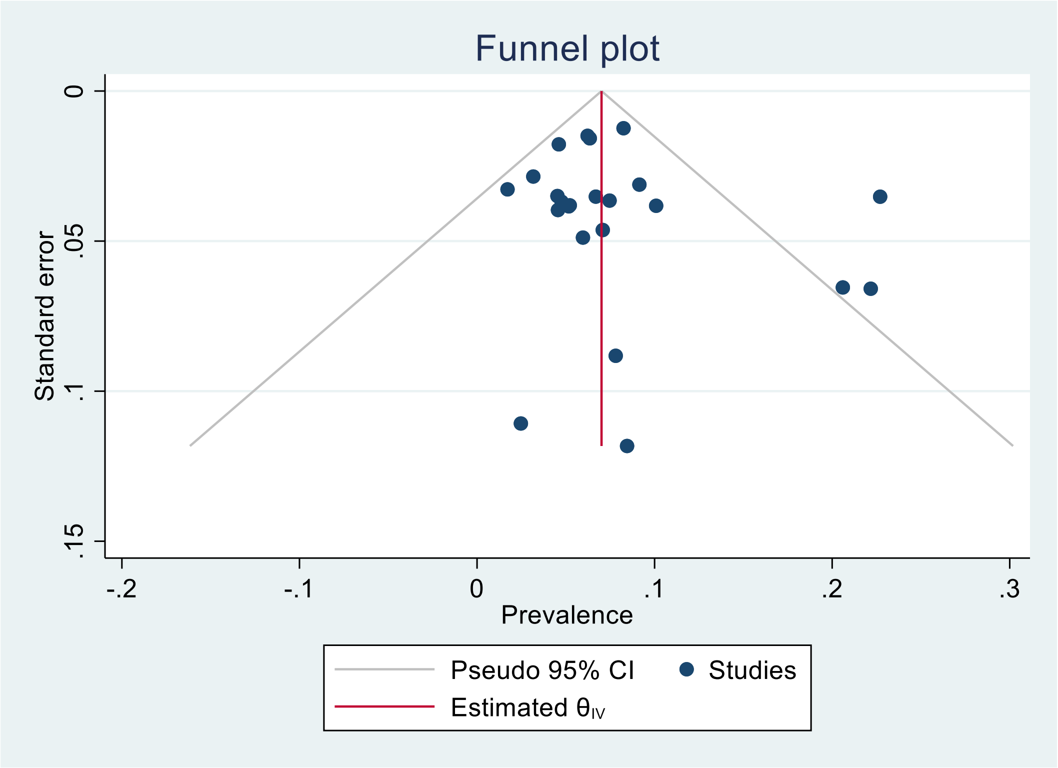


Figure S7: Funnel plot for meta-analysis of the prevalence of all-type dementia for men to same-sex participants.

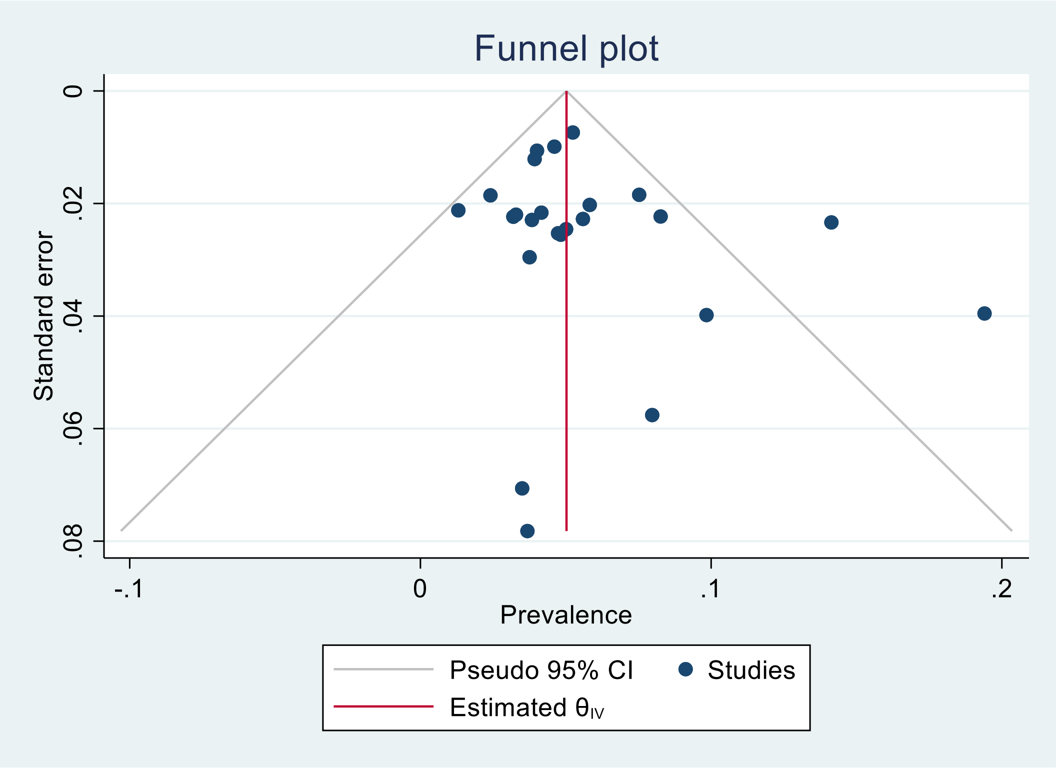


Figure S8: Funnel plot for meta-analysis of the prevalence of all-type dementia for women for the overall participants.

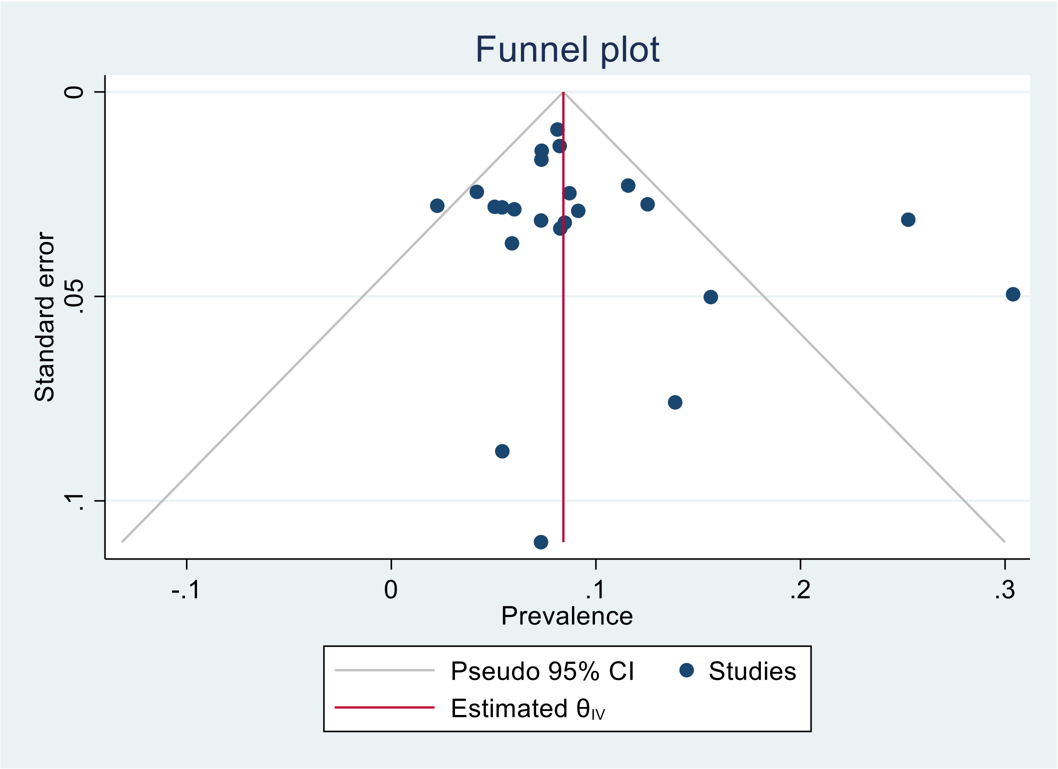


Figure S9: Funnel plot for meta-analysis of the prevalence of all-type dementia for women to the same-sex participants.

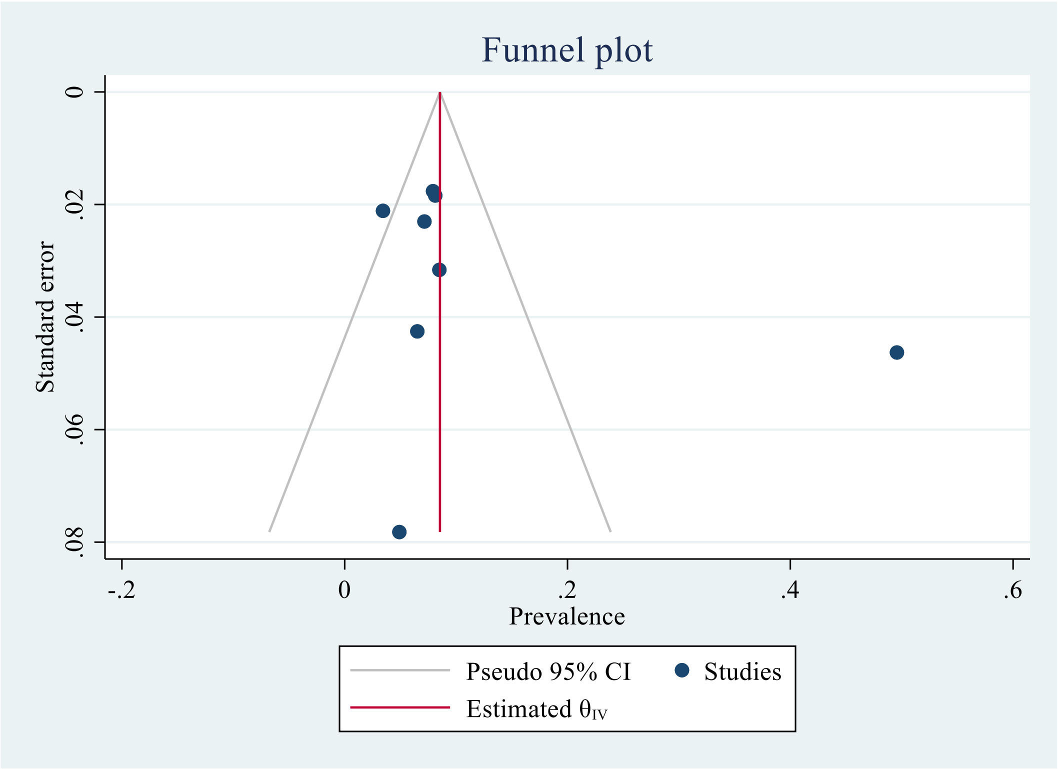


Figure S10: Funnel plot for meta-analysis of the prevalence of all-type dementia for participants from rural area

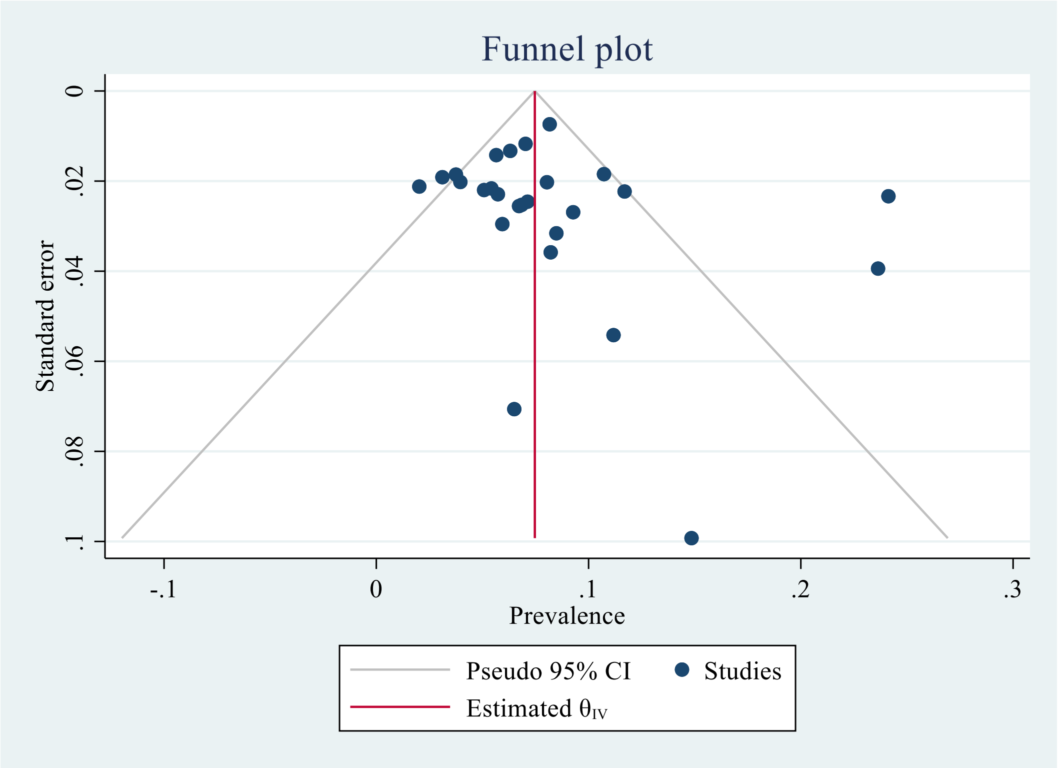


Figure S11: Funnel plot for meta-analysis of the prevalence of all-type dementia for participants from urban area

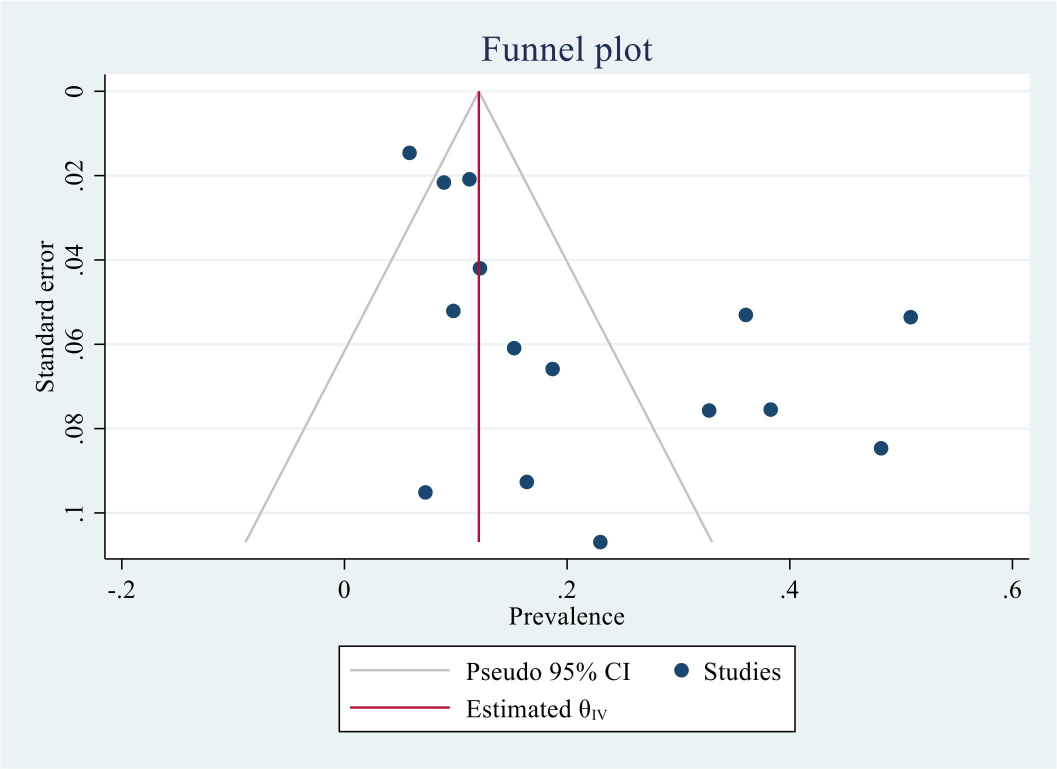


Figure S12: Funnel plot for meta-analysis of the prevalence of all-type dementia for participants with no formal education

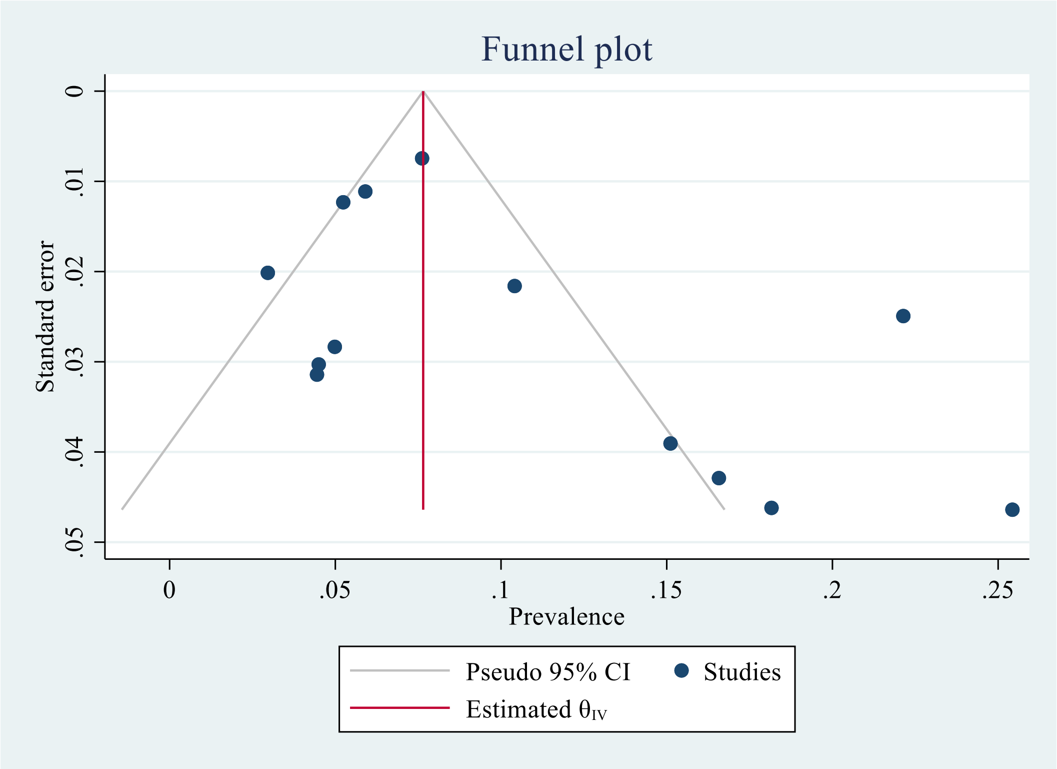


Figure S13: Funnel plot for meta-analysis of the prevalence of all-type dementia for participants with at least one year of formal educatio
